# Efficient count-based models improve power and robustness for large-scale single-cell eQTL mapping

**DOI:** 10.1101/2025.01.18.25320755

**Authors:** Zixuan Eleanor Zhang, Artem Kim, Noah Suboc, Nicholas Mancuso, Steven Gazal

## Abstract

Population-scale single-cell transcriptomic technologies (scRNA-seq) enable characterizing variant effects on gene regulation at the cellular level (e.g., single-cell eQTLs; sc-eQTLs). However, existing sc-eQTL mapping approaches are either not designed for analyzing sparse counts in scRNA-seq data or can become intractable in extremely large datasets. Here, we propose jaxQTL, a flexible and efficient sc-eQTL mapping framework using highly efficient count-based models given pseudobulk data. Using extensive simulations, we demonstrated that jaxQTL with a negative binomial model outperformed other models in identifying sc-eQTLs, while maintaining a calibrated type I error. We applied jaxQTL across 14 cell types of OneK1K scRNA-seq data (*N*=982), and identified 11-16% more eGenes compared with existing approaches, primarily driven by jaxQTL ability to identify lowly expressed eGenes. We observed that fine-mapped sc-eQTLs were further from transcription starting site (TSS) than fine-mapped eQTLs identified in all cells (bulk-eQTLs; *P*=1×10^−4^) and more enriched in cell-type-specific enhancers (*P*=3×10^−10^), suggesting that sc-eQTLs improve our ability to identify distal eQTLs that are missed in bulk tissues. Overall, the genetic effect of fine-mapped sc-eQTLs were largely shared across cell types, with cell-type-specificity increasing with distance to TSS. Lastly, we observed that sc-eQTLs explain more SNP-heritability (*h*^*2*^) than bulk-eQTLs (9.90 ± 0.88% vs. 6.10 ± 0.76% when meta-analyzed across 16 blood and immune-related traits), improving but not closing the missing link between GWAS and eQTLs. As an example, we highlight that sc-eQTLs in T cells (unlike bulk-eQTLs) can successfully nominate *IL6ST* as a candidate gene for rheumatoid arthritis. Overall, jaxQTL provides an efficient and powerful approach using count-based models to identify missing disease-associated eQTLs.

## Introduction

Large gene expression quantitative trait loci (eQTLs) studies have facilitated interpreting genetic variants identified in genome-wide association studies (GWAS) through colocalization ^1–4^ or transcriptome-wide association studies (TWAS) ^5–9^. These approaches have been largely dependent on eQTLs discovered from bulk-RNA sequencing (bulk-eQTLs) on tissue samples ^10,11^ or on a limited number of cell types ^12– 14^. However, limited overlap between bulk-eQTLs and GWAS risk loci ^4,10,15–18^ has hindered the functional interpretation of genetic risk variants and their translation to therapeutic development for human diseases. Multiple hypotheses could explain this “missing link” between GWAS and eQTLs, including the lack of disease-relevant cell types/contexts, cell-type-specific eQTL effects diluted in bulk samples, and limited statistical power to detect weak-effect eQTLs ^15,16^.

Recent and ongoing generation of large scale single-cell RNA sequencing (scRNA-seq) datasets allow direct interrogation of these hypotheses by quantifying gene expression across heterogeneous cell types for a large number of individuals ^19,20^. For example, the OneK1K project has released scRNA-seq data from 1.27 million peripheral blood mononuclear cells (PMBCs) of 982 donors ^19^, with plans to profile 50 million cells in 10,000 donors (TenK10K) ^21^. A current challenge is thus to efficiently identify single-cell (sc-)eQTLs from sparse counts data in these extremely large datasets. Previous sc-eQTL studies ^22– 26^ have leveraged pseudobulk data and used tools that are designed for bulk-eQTL mapping (e.g., Matrix eQTL ^27^, FastQTL ^28^, and tensorQTL ^29^). These tools fit linear models after data normalization on the gene expression matrix ^30,31^. Although the model fitting step is computationally efficient, the eQTL effect on gene expression is less interpretable due to the data transformation (e.g., inverse rank transform). Moreover, for sparse read counts observed in scRNA-seq, transformations are less effective due to sheer number of zeros ^32,33^. Recent studies have proposed modelling the expression of single cells by fitting mixed effect models, either using off-the-shelf R functions ^34^ or under bespoke software such as CellRegMap ^35^ and SAIGE-QTL ^36^. While these approaches improve upon bulk-eQTL mapping approaches, they can become computationally intractable for extremely large single-cell datasets currently being generated at a population scale. In addition to these computational challenges, the characterization of sc-eQTLs across cell types is further complicated by the differential statistical power induced by differences in cell abundances. For example, recent work reported sc-eQTLs were largely cell-type-specific ^19^, in contrast to higher levels of sharing across cell types when eQTLs were identified from sorted RNA-seq data ^37^. Therefore, sc-eQTL mapping and characterization stands to benefit from scalable and statistically powerful software.

To address these limitations, we propose jaxQTL, an efficient software to perform large-scale sc-eQTL mapping using flexible, count-based models. Under simulations, we found that a negative binomial (negbinom) model outperforms linear and Poisson models in identifying sc-eQTLs while maintaining calibrated type I errors. By analyzing OneK1K, we found that jaxQTL with a negative binomial model identifies more eGenes than other models and existing softwares, such as tensorQTL and SAIGE-QTL. Importantly, we found that sc-eQTLs effects were largely consistent across cell types, with cell-type-specificity increasing with distance to transcription start site. Finally, we found that sc-eQTLs explained a greater fraction of heritability for GWAS immune traits compared with bulk-eQTLs, thus improving but not closing the missing link between GWAS and eQTLs. Taken together, our results demonstrate that jaxQTL is a scalable tool in identifying sc-eQTLs by analyzing large single-cell datasets to improve the biological interpretation of genetic risk at disease-relevant cell types.

## Results

### Overview of jaxQTL

We provide a brief overview of jaxQTL model assumptions and inferential pipeline. Given pseudobulk counts *y*_*c*_ for a focal gene in a cellular context c (i.e. summed across all cells of type c), covariates *X*(e.g., age, sex, genotyping principal components), and a cis-genetic variant *g*, jaxQTL implements a generalized linear model (GLM) according to,

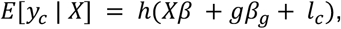

where *β*are the covariate effects, *β*_*g*_ is the allelic effect, *l*_*c*_ is an offset adjusting for differences in library size, and *h*(·) is a function which maps linear predictions to expected values matching distribution assumptions (e.g., negbinom, Poisson). For example, if we assume a Poisson or negative binomial distribution, then *h* := *exp*(·), with effect sizes reflecting a change in the transcription rate (or proportion if including library size offsets). This is in contrast to linear regression performed on the rank-inverse normal transformed counts, where effect sizes have no direct interpretation of the expression values, but rather reflect a change in rankings.

Performing cis-association scans using this approach is computationally prohibitive, due to the sheer number tests required for each gene and cell-type. To address this fundamental limitation, jaxQTL leverages three key insights. First, jaxQTL performs *just-in-time* (JIT) compilation provided by the JAX framework (**Web Resources**), to translate high-level Python into machine-level instructions optimized for a specific parallelized architectures (e.g., CPU, GPU, or TPU) with no additional work required from the user other than a runtime flag. Second, jaxQTL performs a score-test^38^ for all cis-genetic variants simultaneously using an optimized block-matrix approach, rather than sequentially. Lastly, jaxQTL implements multiple recent advances to compute p-values efficiently, which provide trade-offs between additional scalability and statistical power (e.g., Beta-approximation ^28^ to permutations or ACAT-V ^39^; **Figures S1, S2; Methods**). We provide a table summarizing the capabilities of jaxQTL alongside other softwares (**Table S1**) and have released jaxQTL as open-source software (see **Code availability**).

### Negative binomial outperforms other models in identifying sc-eQTLs in realistic simulations

We assessed the type I error and power of different models implemented in jaxQTL (jaxQTL-linear, jaxQTL-negbinom, and jaxQTL-Poisson) and softwares (SAIGE-QTL and tensorQTL) by simulating single-cell read counts from a Poisson mixed effect (PME) generative model using parameters that reflect observed expression and overdispersion in OneK1K ^30,40–46^ (**Figure S3, S4**). We evaluated model performance across varying cell type proportions by sampling individual library sizes across three cell types representing high, medium, and low library sizes (CD4+ naïve and central memory T (CD4_NC_) cells, immature and naïve B (B_IN_) cells, and Plasma cells, respectively; **Table S2**). We varied sample-coverage (i.e., the percentage of non-zero expression read counts) across simulations to account for gene expression intensity across individuals.

First, all models exhibited calibrated type I error rates when simulating from the single -cell PME model (**Figure 1A**), except for pseudobulk jaxQTL-Poisson which displayed increased false positives likely due to its over-conservative standard errors. We observed largely similar conclusions for jaxQTL when varying heritability, random intercept variance *σ*^2^_*u*_(modeling similarity of cell read counts within the same person; see **Methods**), sample size, and minor allele frequency (MAF) parameters (**Figures S5-S8**). Importantly, jaxQTL-negbinom and linear models remain calibrated across cell type abundances and sample sizes, unlike SAIGE-QTL which exhibited increased false positives in rarer cell types when sample sizes are small (N < 200; **Figure S7**). SAIGE-QTL and jaxQTL-negbinom had slight inflation when *σ*^2^_*u*_*≈* 1, however this scenario is unlikely to occur in practice (**Figure S3**).

**Figure 1:**
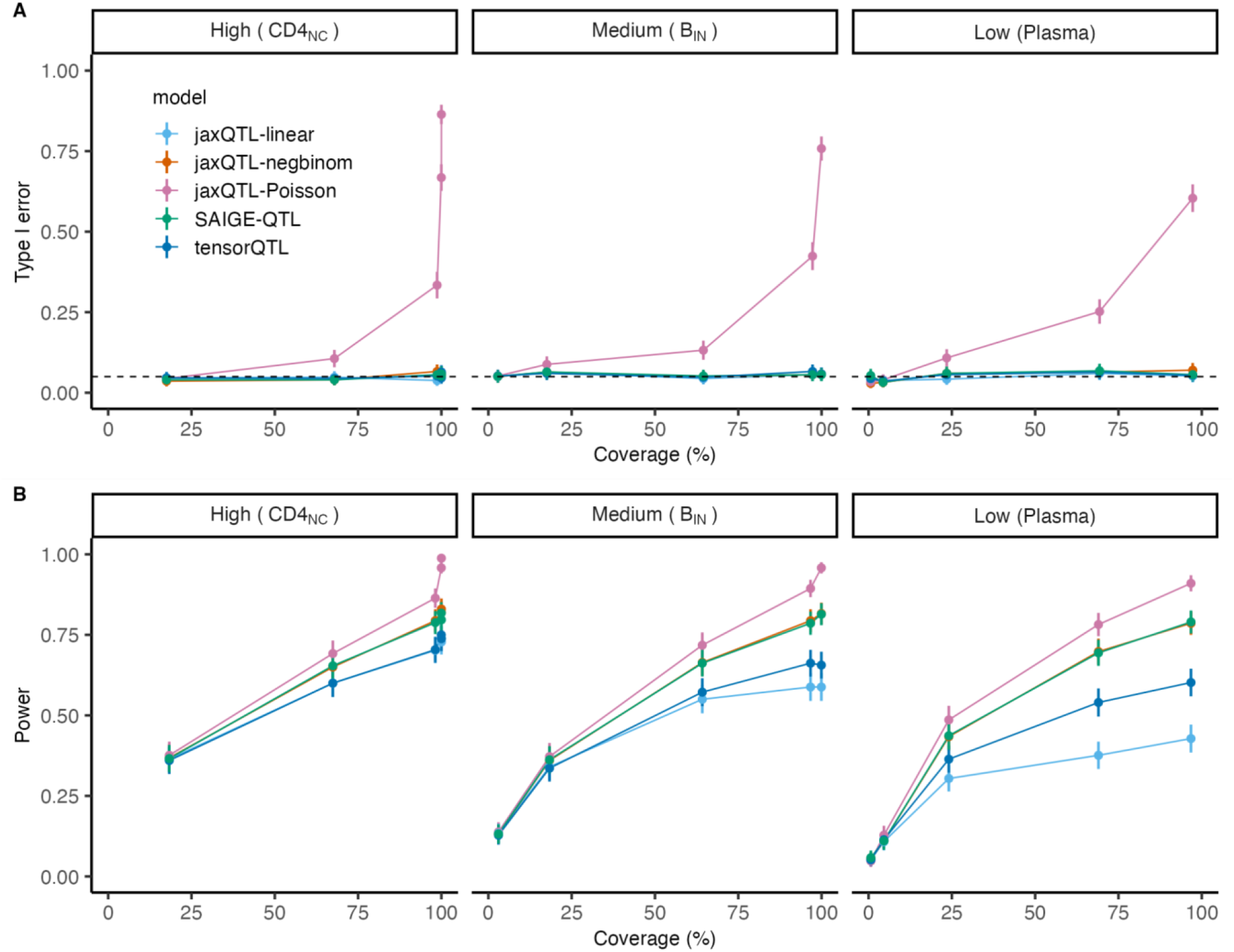
Negative binomial outperforms other models in identifying sc-eQTLs in realistic simulations. We simulated single-cell read counts using library size observed in three cell types (CD4_NC_, B_IN_, Plasma) representing different levels of cell type proportions (high, medium, low). We reported the type I error rate (*h*^2^_*cis*_= 0) **(A)** and power **(B)** of jaxQTL-linear, jaxQTL-negbinom, jaxQTL-Poisson, SAIGE-QTL, and tensorQTL models across different sample-coverage (i.e., percentage of non-zero expression read counts). We fixed cis-heritability *h*^2^_*cis*_= 0.05, random intercept variance *σ*^2^_*u*_(modeling similarity of cell read counts within the same person) = 0.2, sample size = 1,000, and MAF = 0.2; results when varying these parameters are reported in **Figures S5-S8**. Error bars represent 95% confidence intervals (CIs) estimated from 500 replicates. The dashed line in **(A)** represents a type I error of 0.05. SAIGE-QTL assumes single-cell counts while the rest assumes pseudobulk counts.

Next, we observed that jaxQTL-negbinom had improved power compared with jaxQTL-linear and tensorQTL, especially for lower coverage genes. Across three cell types, genes with higher coverage exhibited greater statistical power to identify their sc-eQTLs. Specifically, for large cell type proportions, jaxQTL-negbinom outperformed jaxQTL-linear for genes with >95% coverage (*P* = 7.53 × 10^−4^). For medium and rare cell types, jaxQTL-negbinom exhibited greater power over jaxQTL-linear down to ∼70% coverage (*P* = 2.02 × 10^−4^ and 3.91 × 10^−27^ respectively), highlighting the benefit of count-based models for lower expressed genes and rarer cell types. While both jaxQTL-linear and tensorQTL fit linear models of gene expression, differences in power can be explained by jaxQTL score test versus tensorQTL Wald test. We obtained similar conclusions when varying heritability, random intercept variance *σ*^2^_*u*_, sample size, and MAF parameters (**Figures S5-S8**).

After assessing model performances under the PME model with non-zero random intercept variance *σ*^2^_*u*_, we repeated our analyses by sampling read counts from PME models with *σ*^2^_*u*_= 0, which reflects a standard Poisson model (**Figure S9**). As expected, the performance of jaxQTL-negbinom and SAIGE-QTL closely resembled jaxQTL-Poisson (see **Supplemental Note**). Again, count-based models outperformed jaxQTL-linear and tensorQTL, notably for genes in rarer cell types. All models were well-calibrated under the null.

Altogether, jaxQTL-negbinom provides a calibrated and powerful pseudobulk model for single-cell data that performs comparably to the PME model of SAIGE-QTL. Our empirical results can be in part explained by the structural similarity of the variance under negative binomial and PME models of pseudobulk (see **Supplemental Note**).

### jaxQTL improves power for eGene discovery in the OneK1K dataset

To benchmark jaxQTL in identifying eGenes on real datasets, we applied jaxQTL on single-cell data of 14 PBMC cell types from *N* = 982 individuals in OneK1K ^19^. We defined eGene as genes with at least one sc-eQTL in a cell type, i.e., gene-cell-type pairs. Before comparing different sc-eQTL models, we investigated the calibration of gene-level *P* values obtained by the Beta-approximation approach that permutes the gene expressions observed in OneK1K data. Across the three representative cell types, we found that gene-level *P* values from jaxQTL-linear and jaxQTL-negbinom were well-calibrated (**Figure S10**), however gene-level *P* values for jaxQTL-Poisson were inflated due to overcorrection by the permutation method on its variant *P* values.

After confirming the gene-level *P* values were calibrated, we compared the statistical power in identifying eGenes across different models using jaxQTL (**Figure 2A; Table S3**). Across 14 cell types, jaxQTL-negbinom identified 14% more eGenes compared with jaxQTL-linear (18,907 vs. 16,654 eGenes, *P* = 1 × 10^−35^), and 21% more compared with jaxQTL-Poisson (15,634 eGenes, *P* = 5 × 10^−75^). The number of eGenes found per cell type was highly correlated with cell type proportions, which reflects differential statistical power (Pearson *ρ* = 0.97; **Figure S11**). Focusing on jaxQTL-negbinom and jaxQTL-linear, we found the negbinom model provided higher *χ*^2^ test statistics for lead SNP-eGene pairs across cell types (median *χ*^2^ = 43.60 vs. 38.46, *P* = 3 × 10^−24^; **Figure S12**). eGenes identified between models show substantial overlap (**Figure S13A**). Consistent with simulation results, eGenes identified exclusively by jaxQTL-negbinom had lower coverage (median 79% vs. 94%, *P* = 2 × 10^−115^; **Figure S13B**) than eGenes also identified with jaxQTL-linear, confirming that the negbinom model is more powerful for genes with lower expression. The reduced power of the jaxQTL-Poisson was caused by the penalty on its inflated type I error when computing the gene-level *P* values using the permutation approach. Given the improvement of the negbinom model over Poisson and linear models, to simplify our presentation we refer to jaxQTL-negbinom as jaxQTL for the remainder of the manuscript.

**Figure 2:**
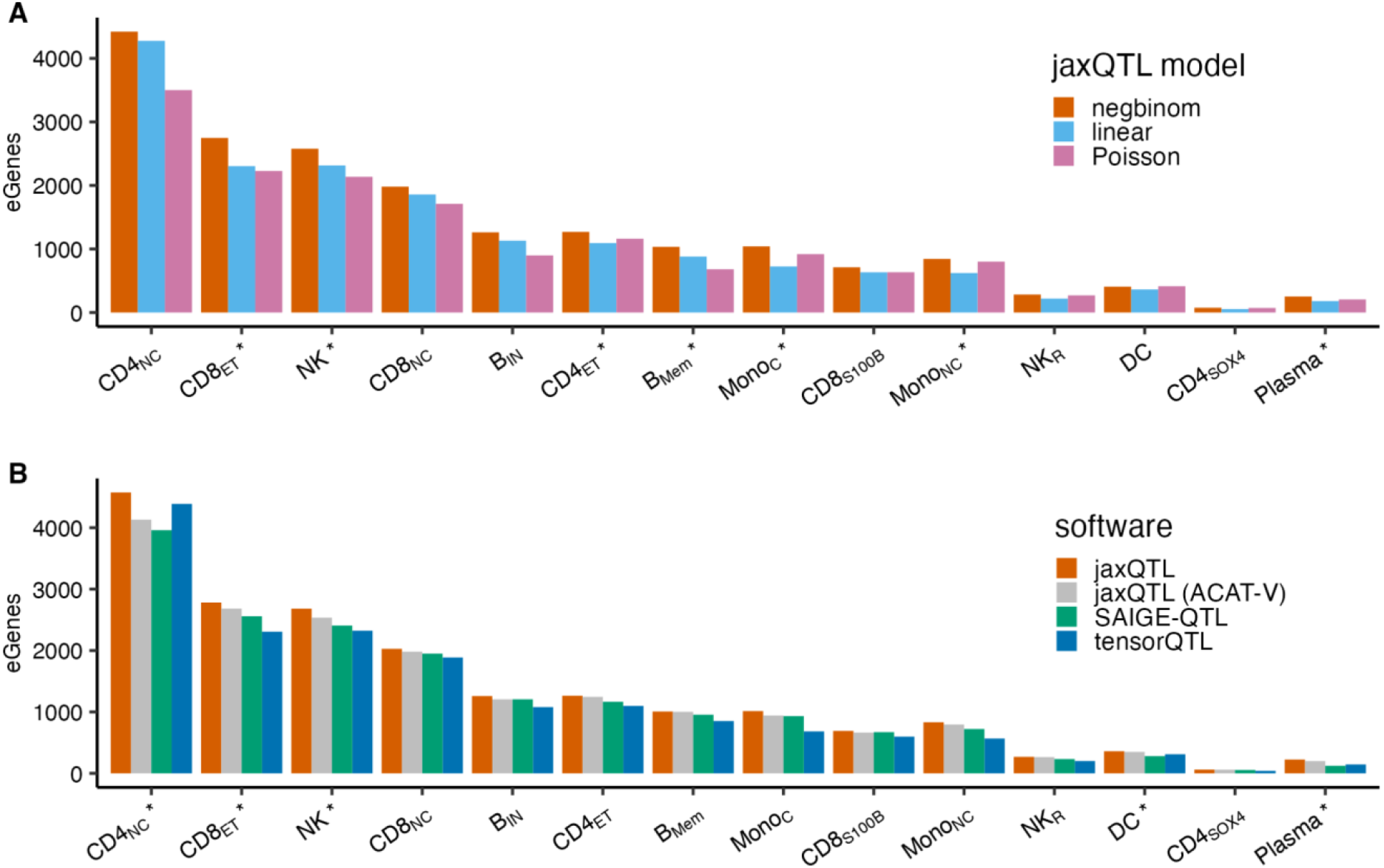
jaxQTL improves power for eGene discovery in the OneK1K dataset. We compared eGene findings in OneK1K across models and software at FDR < 0.05. **(A)** For model comparison, we reported the number of eGenes identified by jaxQTL-negbinom, jaxQTL-linear, and jaxQTL-Poisson for genes with sample-coverage > 1%. **(B)** For software comparison, we reported the number of eGenes identified by jaxQTL (negbinom), jaxQTL (use ACAT-V instead of permutation method), tensorQTL, and SAIGE-QTL for genes with sample-coverage > 10% of individuals. The asterisks denote the cell type in which jaxQTL (negbinom) identified more eGenes compared with the linear model in **(A)** or SAIGE-QTL in **(B)** after Bonferroni correction. See details of description on cell types in **Table S2**. Numerical results are reported in **Table S3, S4**.

We next compared jaxQTL performance against tensorQTL ^29^ (a commonly used software for bulk-eQTL mapping using a linear model) and SAIGE-QTL ^36^ (a recent software for sc-eQTL mapping using a Poisson mixed-effect model) (**Figure 2B; Table S4**). Across 14 cell types, jaxQTL identified 16% more eGenes compared with tensorQTL (*P* = 2 × 10^−47^) and 11% more eGenes compared with SAIGE-QTL (*P* = 3 × 10^−24^), thus demonstrating jaxQTL increased power to identify eGenes in both rare and common cell types. The advantage of jaxQTL over SAIGE-QTL can be partially explained by gene-level calibration methods (permutation vs. ACAT-V), as performance gap decreased when applying ACAT-V in jaxQTL (5% more eGenes; **Figure 2B**). We note that our tensorQTL results identified more eGenes than tensorQTL in ref. ^36^, likely due to different procedures to create pseudobulk data (see **Methods**). Finally, we confirmed that jaxQTL-linear results agreed with our tensorQTL results since the Wald test and score test are asymptotically equivalent (97% overlap; **Figure S14**), with differences up to gene-level *P* value obtained by the permutation approach.

Next, we evaluated the computational performance of jaxQTL in cis-eQTL mapping compared with existing approaches using 50 randomly selected genes from chromosome 1 in OneK1K (see **Methods**; **Figure S15**). The average run time of jaxQTL on GPU/TPU across 3 cell types is 3.7x faster compared with SAIGE-QTL (12 vs. 44 mins) and 9.2x slower compared with tensorQTL (1.3 mins). To demonstrate the impact of sample size on run time, we simulated data for varying sample size by downsampling from N=100 to 700. The average run time is 39 mins for SAIGE-QTL, 15 mins for jaxQTL on GPU/TPU, and 2 mins for tensorQTL. To mimic TenK10K data^21^, we performed upsampling to simulate data for N=10,000 (see **Methods**). Focusing on a dominant cell type such as CD4_NC_ cells, jaxQTL on GPU was at least 1,560x faster (30 mins) compared with SAIGE-QTL, highlighting the efficiency of jaxQTL when applied to ever-increasing population-scale single-cell data. We note that the runtime of jaxQTL is dominated by performing permutations. Importantly, when performing ACAT-V to compute gene-level P values on CPU, jaxQTL was 1.3 - 10,596x times faster than SAIGE-QTL for N=100 to 10,000, and comparable with the linear model of tensorQTL (with permutations).

In summary, jaxQTL outperforms other models and methods in identifying eGenes in OneK1K, highlighting that pseudobulk-approaches for scRNA-seq are powerful (even for rarer cell types) when appropriately modeling count data. In addition, we observed that its computation time can scale to scRNA-seq datasets with thousands of individuals approaching that of classical linear models.

### jaxQTL results replicate across datasets and ancestries

To verify that increased eGene detection from jaxQTL is not driven by false positives, we first replicated our sc-eQTLs results in 88 European- and 88 Asian-ancestry individuals from CLUES PBMC scRNA-seq study (**Figure 3**; **Figure S16, S17; Table S5**)^47^. Of the lead SNP-eGene pairs found in matched CLUES cell types, 40-86% can be replicated in a European cohort and 23-74% in an Asian cohort at FDR < 0.05 with concordant directional effect. Additionally, we replicated 75-92% sc-eQTLs in EUR whole blood samples from GTEx (N=588)^10^ and 50%-78% in FACS-sorted immune cell types from DICE study (N=91)^48^ (**Figure S18; Table S6**). We also observed consistent direction of sc-eQTL effects when comparing with shared lead SNP-eGene results in original OneK1K results (**Figure S19**). Lastly, we recapitulated the depletion of selection constraint and short enhancer domains in eGenes ^10,49,50^ (**Figure S20;** see **Web resources**). Consistent with refs. ^49,51–53^, we observed genes depleted of loss-of-function mutations (pLI > 0.9) had smaller sc-eQTL effect sizes (**Figure S21**), and that the effect size of lead SNPs of eGenes was smaller at lower allele frequency (**Figure S22**), confirming selection constraints on gene expressions in immune cell types. Altogether, these results demonstrate that jaxQTL results replicate across datasets and recapitulate known findings from bulk-eQTL studies.

**Figure 3:**
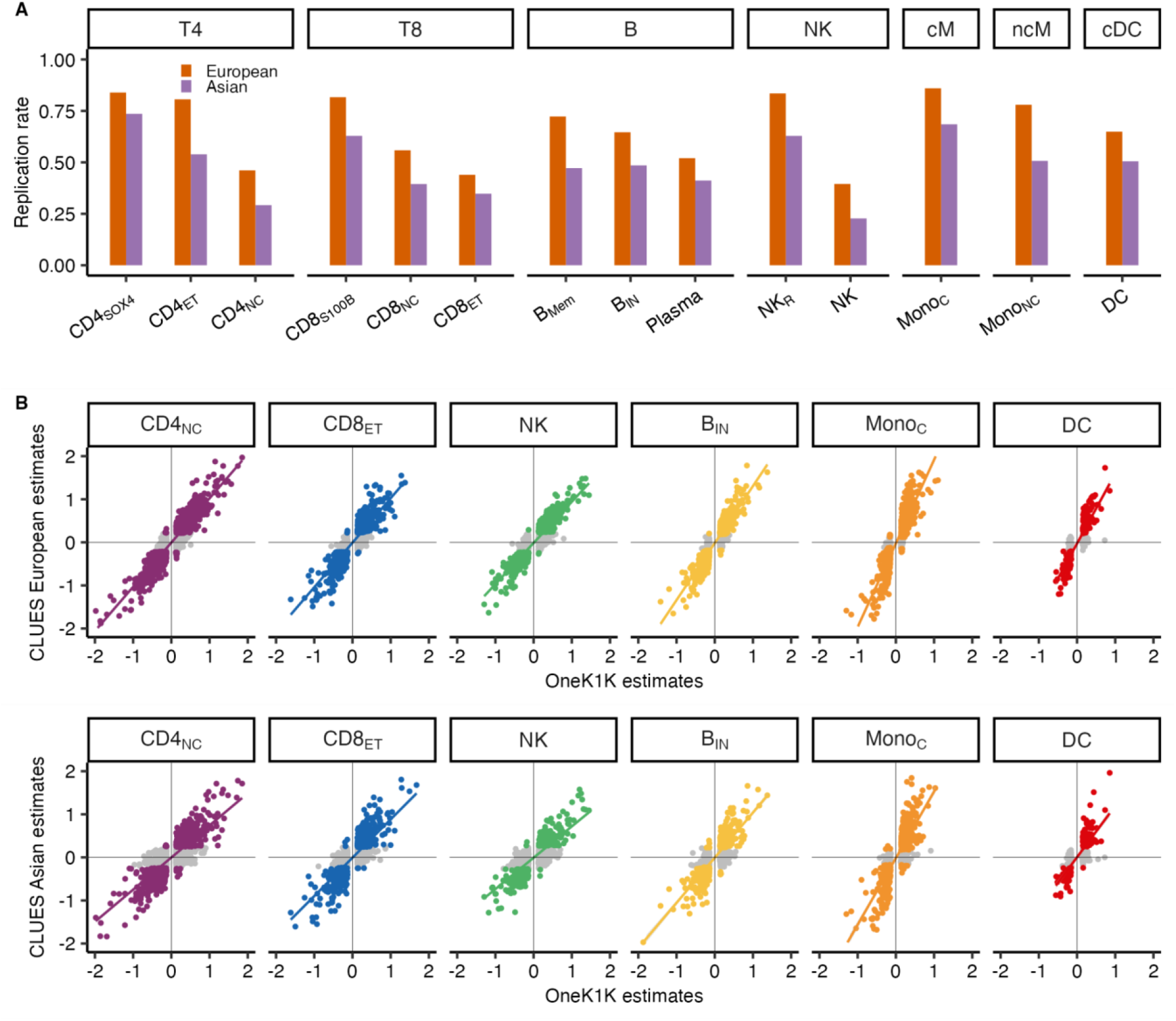
jaxQTL sc-eQTLs replicate in European and Asian samples. We performed replication analysis for 18,907 sc-eQTLs identified by jaxQTL in CLUES study. **(A)** Of the lead SNP-eGene pairs found in matched cell types (panels) among 88 European- and 88 Asian-ancestry individuals (14,229 and 13,579 sc-eQTLs respectively), we reported the replication rate at FDR < 0.05 by ancestry. **(B)** We plotted the adjusted sc-eQTL effect estimated by jaxQTL in CLUES versus OneK1K samples (see **Methods**). Colored points are pairs replicated at FDR < 0.05 in CLUES samples and grey points are otherwise. 35 and 20 pairs with absolute adjusted sc-eQTL effect estimate > 2 were truncated for visualization (see complete results in **Figure S16, S17** and **Table S5**). The colored line in **(B)** is a fitted linear regression line with a 95% confidence band. T4: CD4+ T cells; T8: CD8+ T cells; NK: natural killer cells; cM: CD14+ conventional monocytes; ncM: CD16+ unconventional monocytes; cDC: conventional dendritic cells.

### sc-eQTLs are more enriched in cell-type-matched CREs than bulk-eQTLs

To characterize sc-eQTLs and their potential downstream role in human diseases, we performed fine-mapping for eGenes identified by jaxQTL on OneK1K using the SuSiE summary statistics approach ^54^. Briefly, SuSiE performs Bayesian variable selection to identify likely causal SNPs in the form of credible sets and provide posterior inclusion probabilities (PIPs) to quantify uncertainty in its selection (see **Methods**). After restricting to the 18,281 non-MHC and non-MAPT eGenes identified by jaxQTL, SuSiE reported 95% credible sets (CS) for 12,978 eGenes across 14 cell types (6,776 unique eGenes). The average number of CSs per eGene was 1.15 with a median size of 16 SNPs, with 88% of eGenes explained by a single causal variant. We observed that the average number of CSs tracked with cell type proportion (*P* = 1.49 × 10^−5^; **Figure S23**), suggesting the fine-mapping results were likely biased by lower statistical power to pinpoint independent causal eQTLs in rarer cell types. To establish a baseline, we also performed cis-eQTL and fine-mapping analyses using “bulk” gene expression (i.e., summed over cell types). As expected, we found that sc-eQTLs successfully identified a CS for more unique eGenes than bulk-eQTLs (6,776 vs 6,338, *P* = 9.5 × 10^−23^).

To characterize the functional architecture of fine-mapped sc-eQTLs, we performed enrichment analysis using cell-type-agnostic annotations from S-LDSC baseline model^55^ (see **Methods**). Consistent with bulk-eQTLs ^56^, fine-mapped sc-eQTLs (PIP ≥ 0.5) across cell types were highly enriched in promoter-like regions, enhancers, and evolutionarily conserved regions (**Figure 4A**). Overall, we observed greater enrichments for these annotations when using sc-eQTLs compared with bulk-eQTLs, however these differences were not significant, likely resulting from baseline annotations not reflecting cell-type-specificity. Additionally, we observed fine-mapped sc-eQTLs were less likely to be near promoter regions (*P* = 6.71 × 10^−4^) and more distal (*P* = 1.07 × 10^−4^) when compared with bulk-eQTLs (**Figure 4B**), suggesting that single-cell eQTL mapping can better prioritize distal regulatory elements.

**Figure 4:**
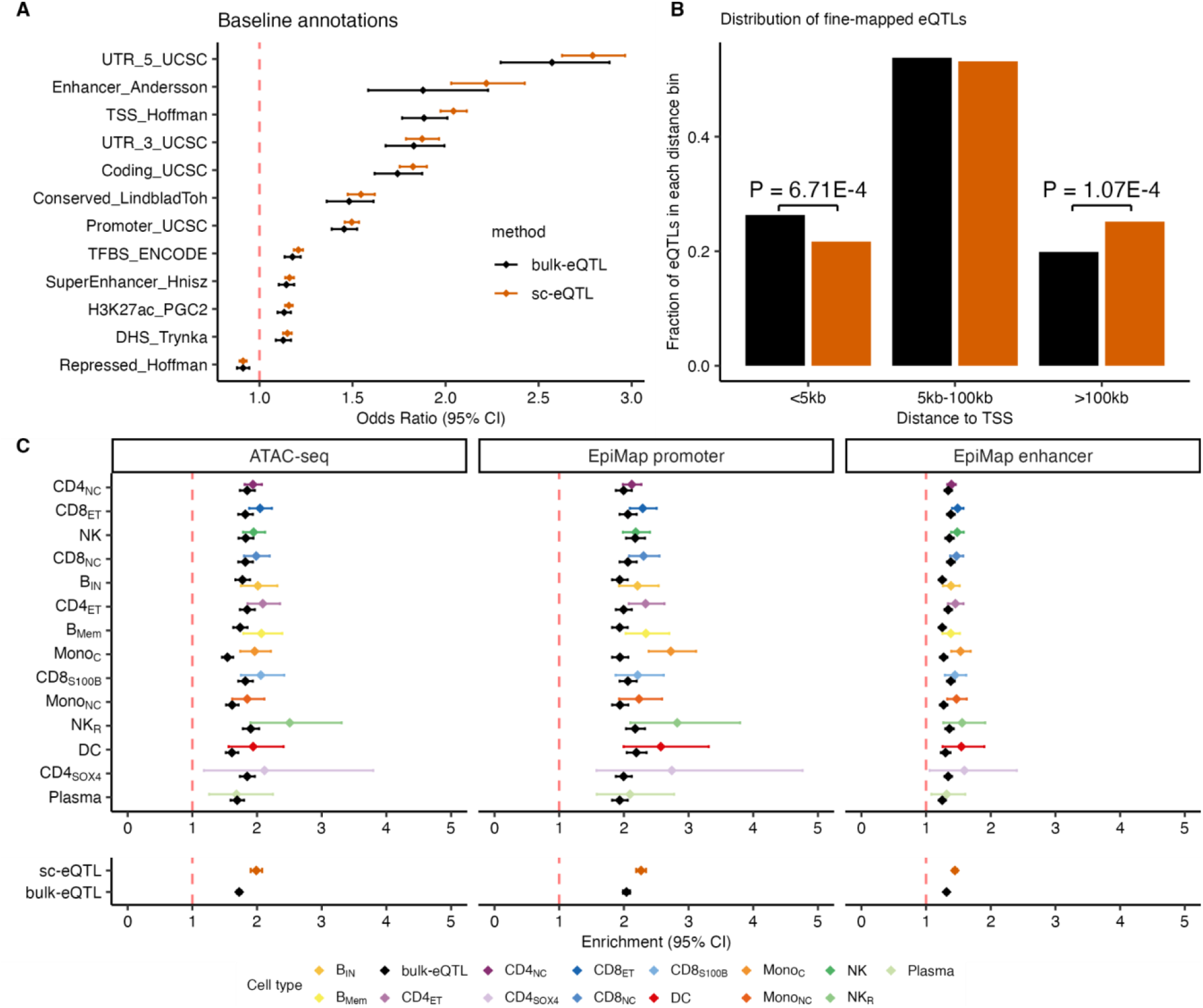
sc-eQTLs are more enriched in cell-type-matched CREs than bulk-eQTLs. We performed enrichment analysis of sc-eQTLs and bulk-eQTLs using their fine-mapping results and diverse annotations. **(A)** We report the odds ratio for eQTLs enrichment within 12 S-LDSC representative baseline annotations (see **Methods**). **(B)** For fine-mapped sc-eQTLs with PIP≥0.5, we report the fraction of fine-mapped eQTLs falling in three distance to TSS bins. **(C)** We report the enrichment in sc-eQTLs per cell type and bulk-eQTLs within 3 types of cell-type-specific functional annotations; we report meta-analysis results across 14 cell types at the bottom of each dataset. Error bars represent 95% CIs. Numerical results are reported in **Tables S11-14**.

Next, we sought to compare the enrichment of cell-type-specific candidate cis-regulatory elements (cCREs), derived using single-cell omics or isolated cell types, between fine-mapped sc-eQTLs and bulk-eQTLs. First, we confirmed that sc-eQTLs were enriched in cell-type-matched cCREs reflecting open chromatin, enhancers, and promoters (**Figure 4C; Methods**). We observed that sc-eQTLs were highly enriched in enhancer-gene links in matched cell types. Importantly, sc-eQTLs were more enriched in cell-type matched open chromatin (*P* = 6.77 × 10^−9^), enhancers (*P* = 3.40 × 10^−10^) and promoters (*P* = 1.26 × 10^−6^) compared with bulk-eQTLs after meta-analysis across cell types (**Figure 4C**). Lastly, we further confirmed the accuracy of sc-eQTLs linked to their target genes by observing stronger enrichment in cell-type-matched enhancer-gene links identified by SCENT ^57^ compared to bulk-eQTLs (*P* = 3.38 × 10^−4^; **Figure S24**), highlighting the necessity of conducting eQTL mapping using single-cell data to capture signals masked by bulk approach.

In summary, our analyses suggest that sc-eQTLs are enriched for distal cell-type-specific CREs that are likely missed by bulk-eQTL approach.

### sc-eQTL location predicts cell-type specificity

Understanding how sc-eQTLs are shared across cell types is challenging due to differential statistical power between cell types. Specifically, simply counting sc-eQTLs based on their significance or fine-mapping results would conclude to a high cell-type specificity of sc-eQTLs (**Figure S25**; results similar to the ones reported by ref. ^19^), but ignores the pervasive correlation of eQTL effects between cell types (**Figure S26**; as observed using eQTLs from bulk cell type samples in ref. ^37^). To mitigate this, we analyzed fine-mapped sc-eQTLs using *mashr*, which provides posterior effect estimates and significance of effect after accounting for the effect size correlation between cell types and residual correlation due to sample overlap ^58^ (2,012 sc-eQTLs with PIP ≥ 0.5 and significant at a local false sign rate (LFSR) < 0.05 in at least one cell type).

Using the *mashr* estimated effect sizes, we found that 67% of fine-mapped sc-eQTLs were shared by sign across all 14 cell types (**Figure 5A**), suggesting their directional effect on gene expression was consistent. In contrast, eQTL effect size magnitudes were less shared due to effect size heterogeneity, with 15% universally shared and 9% specific (**Figure 5A**), consistent with previous findings ^10,58,59^. Cell-type-specific sc-eQTLs were most common in monocytes, reflecting their difference from lymphocytes such as B cells and T cells (**Figure S27**).

**Figure 5:**
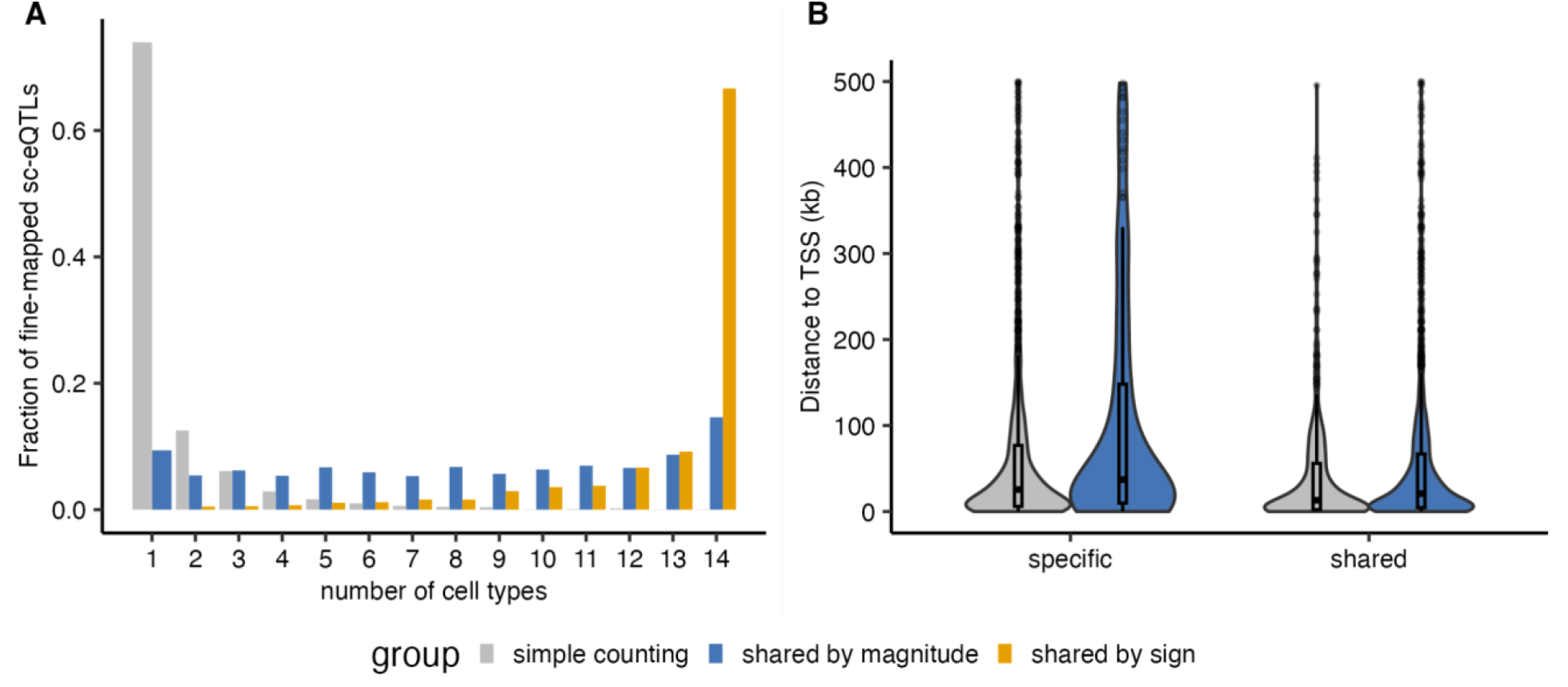
sc-eQTL location predicts cell-type specificity. We investigated sc-eQTL sharing across cell types by performing *mashr* analysis ^58^ on 2,012 OneK1K sc-eQTLs fine-mapped in at least one cell type; as a baseline, we also investigated sc-eQTL sharing by simple counting approach. **(A)** We report the fraction of sc-eQTL shared across different numbers of cell types using three approaches: a simple counting approach, *mashr* estimates of sharing by magnitude (i.e., the magnitude of effect size is within a factor of 2 compared to the strongest signal), *mashr* estimates of sharing by sign (i.e. the sign of effect size is shared with this discovery cell type, i.e., the cell type with the strongest signal). **(B)** We report the distance to TSS for sc-eQTLs identified as cell-type-specific or shared in at least 2 cell types using a simple counting approach and *mashr*. The median value of distances is displayed as a band inside each box; boxes denote values in the second and third quartiles; the length of each whisker is 1.5 times the interquartile range, defined as the height of each box. Numerical results are reported in **Tables S15**.

We found cell-type-specific sc-eQTLs identified were more distal from TSS compared with shared sc-eQTLs (mean 102,164 vs. 53,733 bp, *P* = 2.17 × 10^−6^; **Figure 5B**), consistent with previous work showing that distal CREs were more likely to be cell-type-specific ^60,61^. Lastly, we calculated the pairwise sharing of sc-eQTL by magnitude and found cell type sharing outside expected subtype groups (**Figure S28**). For example, we found 84% shared sc-eQTLs between effector memory CD8+ T cells and natural killer (NK) cells, suggesting their shared cytotoxic effector mechanisms between adaptive and innate immunity ^62^.

To validate the specificity of cell-type-specific sc-eQTLs identified by *mashr* (in comparison with a baseline simple counting approach), we first calculated the replication rate and observed that cell-type-specific eQTLs after *mashr* analysis were less replicated in eQTLGen (0.60 vs. 0.77, *P* = 3.11 × 10^−7^) and GTEx (0.33 vs. 0.47, *P* = 5.19 × 10^−4^) respectively, consistent with the expectation that sc-eQTL effects private to a single cell type were likely masked by the bulk-eQTL study. Second, we found the cell-type-specific sc-eQTLs identified by *mashr* were more distal from TSS compared to the simple counting approach (mean 102,164 vs. 63,873 bp, *P* = 9.56 × 10^−4^), suggesting *mashr* was more powerful in identifying distal cell-type-specific sc-eQTLs. Finally, we identified scATAC-seq peaks exclusive to each cell type and calculated the enrichment of cell-type-specific open chromatins. We observed the specific eQTLs identified by *mashr* were more enriched in cell-type-specific open chromatin in rarer cell types (**Figure S29**), such as non-classical monocytes (Mono_NC_), NK recruiting (NK_R_) cells, CD4+ T cells expressing SOX4 (CD4_SOX4_), and Plasma, which reflected eQTL sharing results (**Figure S27**).

In summary, our analyses suggest that the genetic effect of sc-eQTLs are largely shared across cell types. sc-eQTLs closer to TSS are more likely to be shared while distal sc-eQTLs are likely cell-type-specific due to the different regulatory elements involved in transcription.

### sc-eQTLs reveal cell types associated with GWAS immune traits

After observing that sc-eQTLs improved our ability to identify cell-type-specific eQTLs, we next investigated whether sc-eQTLs can improve the interpretation of GWAS findings, which SNP-heritability (*h*^*2*^) tend to be concentrated in SNPs within cCREs ^55,63,64^ and genes active in disease-relevant cell types ^65–67^. To quantify the extent to which sc-eQTLs can characterize GWAS findings, we first evaluated the fraction of *h*^*2*^ explained by SNP-annotations constructed from fine-mapped sc-eQTLs in all PBMC cell types and meta-analyzed S-LDSC results across 16 immune diseases and blood traits (**Table S7**). We found that the union of fine-mapped sc-eQTLs across 14 cell types (3.5% of common SNPs) were enriched in *h*^*2*^ (2.82 ± 0.25), and explained 9.90 ± 0.88% of *h*^*2*^. Importantly, fine-mapped sc-eQTLs explained more *h*^*2*^ than bulk-eQTLs (6.10 ± 0.76%; **Figure 6A**) while maintaining comparable *h*^*2*^ enrichment (2.76 ± 0.34; **Figure S30**), suggesting that sc-eQTLs increase the number of GWAS variants that can be functionally characterized.

**Figure 6:**
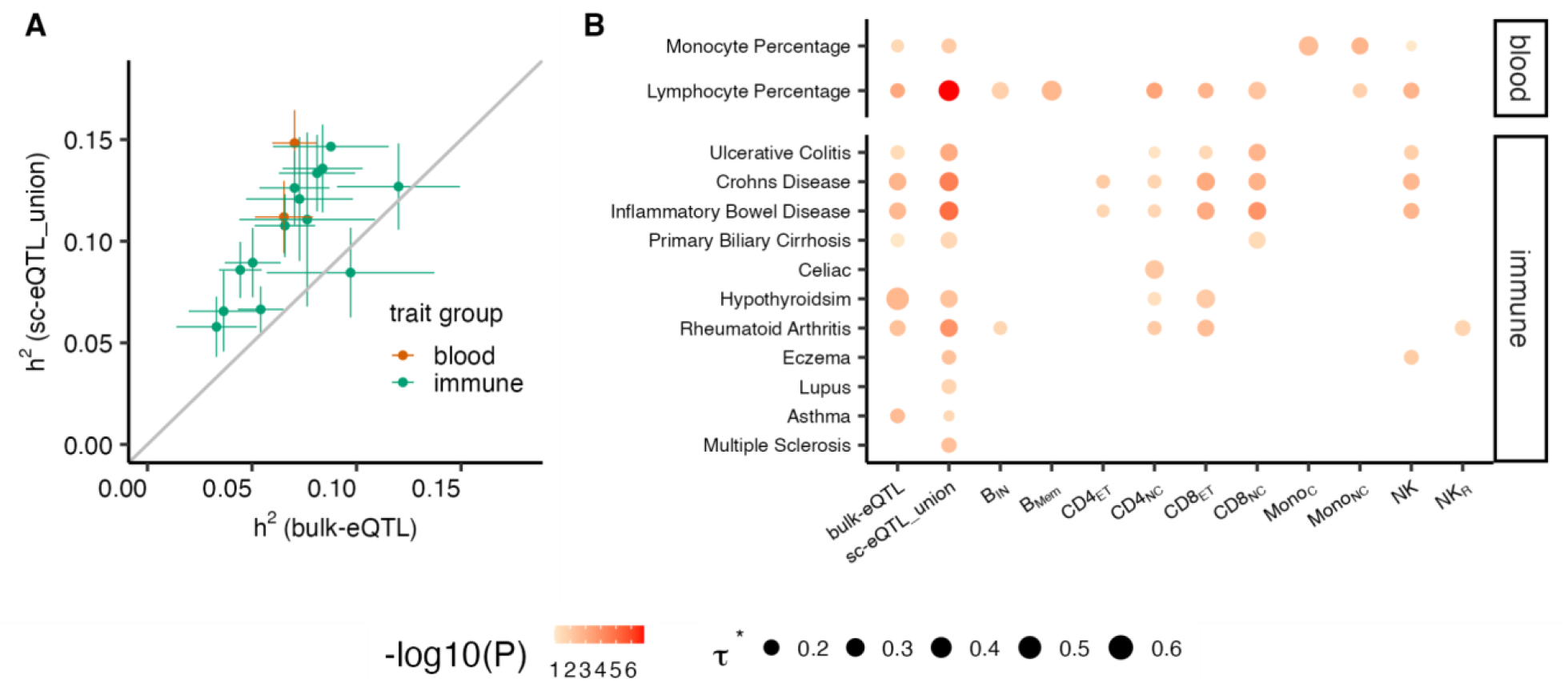
sc-eQTLs explain more heritability than bulk-eQTLs for immune-related GWAS traits. **(A)** We report the proportion of heritability (*h*^*2*^) explained by SNP-annotations built from the union of sc-eQTLs and bulk-eQTLs for 16 GWAS blood and immune-related diseases. Error bars denote 1 standard error of the corresponding estimates. **(B)** We report S-LDSC standardized effect size (*τ*^*\**^) and its associated *P* values obtained for 13 traits and 12 eQTL annotations; *τ*^*\**^represents the proportionate change in per-SNP *h*^*2*^ associated with 1 standard deviation change of annotation value after conditioning to baseline SNP-annotations. Only trait-annotation pairs with significant *τ*^*\**^(after FDR correction) were plotted. The size of the dot is proportional to the standardized effect size *τ*^*\**^, and the colorness of the dot is proportional to -log_10_(*P*). Numerical results are reported in **Tables S16, S17**.

We further investigated whether *h*^*2*^ was enriched within sc-eQTLs from disease-relevant cell types (**Figure 6B**). We ran S-LDSC on sc-eQTL annotations built from each cell type, and individually looked at their effects while conditioning on the union of fine-mapped sc-eQTLs. Overall, identified cell types were consistent with known biology and previous studies leveraging cCREs and genes differentially expressed. For example, we identified monocyte cell types for monocyte percentage (min *P* = 1.80 × 10^−3^ for Mono_NC_) and major sub-cell-types of B cells and T cells for lymphocyte percentage (min *P* = 4.26 × 10^−4^ for CD4_NC_). For immune diseases, we identified various T cell subtypes for celiac disease ^66^ (min *P* = 7.65 × 10^−3^ for CD4_NC_), inflammatory bowel disease ^66^ (min *P* = 1.34 × 10^−4^ for naïve and central memory CD8+ T cells (CD8_NC_)), hypothyroidism ^67^ (min *P* = 9.91 × 10^−3^ for CD8_ET_), primary biliary cirrhosis ^66^ (min *P* = 3.45 × 10^−2^ for CD8_NC_), and rheumatoid arthritis ^66,68^ (min *P* = 3.43 × 10^−3^ for CD8_ET_), as well as the role of NK cells for eczema ^69^(min *P* = 1.12 × 10^−2^ for NK).

Altogether, these results highlight that sc-eQTLs can improve our ability to characterize GWAS findings by identifying new eQTLs in disease relevant cell types.

### sc-eQTLs prioritize candidate genes missed by bulk-eQTLs

To demonstrate that sc-eQTLs can prioritize GWAS candidate genes that would have been missed by bulk sequencing, we present OneK1K sc-eQTLs and bulk-eQTLs results at a leading genetic risk loci associated with rheumatoid arthritis (RA) ^70^ in *ANKRD55-IL6ST* region ^71,72^ (**Figure 7**). We identified the GWAS leading SNP rs7731626 (chr5:55444683:G>A) as the top candidate causal sc-eQTLs for *IL6ST* in CD4_NC_ (PIP = 1) and CD8_NC_ T cells (PIP = 0.98), and observed significant colocalization with RA for these two cell types (PP.H4 = 1, and PP.H4 = 0.99, respectively). In contrast, this SNP became null in bulk-eQTL results as its eQTL effect on *IL6ST* was diluted when cell types were lumped together. We further confirmed that rs7731626 is likely to be within an enhancer acting on *IL6ST* regulation in T cells by observing contact between rs7731626 and *IL6ST* promoter exclusively in T cells using promoter capture Hi-C (PCHi-C) experimental data ^73^, as well as H3K27ac peaks (capturing enhancer activity) at this locus in naive CD4+ and CD8+ T cells ^74,75^ (**Figure 7**; see **Code and Data Availability**); the link between rs7731626 and *IL6ST* in T cells has also been established by other single-cell multi-omic data approaches ^34,76–78^. We replicated the rs7731626-*IL6ST* association in CD4+ and CD8+ T cells in CLUES European- and Asian-ancestry individuals (N=88, p=0.03 respectively), except rs7731626 in CD8+ T cells among Europeans (p=0.3), likely due to lower sample size for detecting weaker effect as evidenced by consistent effect direction. Besides, we also identified rs7731626 as both a bulk-eQTL and sc-eQTL for *ANKRD55* and similar colocalization with RA (**Figure S31**), illustrating that while bulk-eQTL approaches can identify strong sc-eQTL signal, they can nominate an incomplete list of candidate genes.

**Figure 7:**
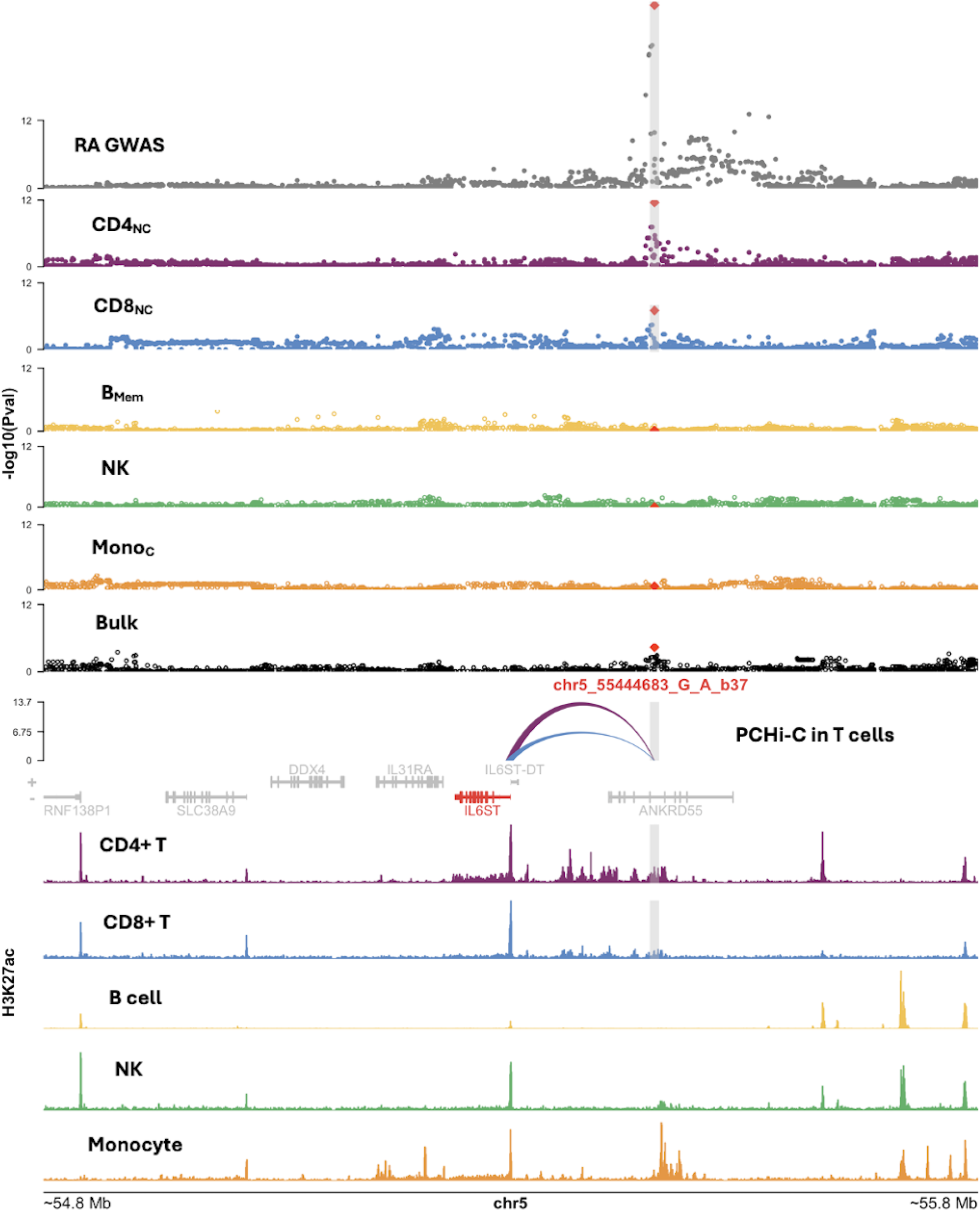
sc-eQTLs prioritize candidate genes missed by bulk-eQTLs. We present an example showing that OneK1K sc-eQTLs in CD4_NC_ and CD8_NC_ can nominate *IL6ST* as a candidate gene for RA, while OneK1K bulk-eQTLs cannot. We report RA GWAS results at the locus highlighting GWAS leading SNP rs7731626 (chr5:55444683:G>A) (1^st^ row), significant *IL6ST* sc-eQTLs in CD4_NC_ and CD8_NC_ T cells (2^nd^ and 3^rd^ row), non-significant (represented by open circle) sc-eQTLs in B_Mem_, NK and Mono_C_ cells (4^th^ to 6^th^ row), non-significant bulk-eQTLs (7^th^ row), PCHi-C links ^73^ between rs7731626 loci and TSS of *IL6ST* observed in CD4+ and CD8+ T cells (height of the arch is proportional to the the score of the link; 8^th^ row), and H3K27ac peaks observed in ENCODE samples corresponding to 5 cell types ^74,75^ (height of the bar is proportional to the peak intensity; 9^th^ to 13^th^ row). In Manhattan plots, we report -log_10_(*P*) of all SNPs within ±500kb from TSS of the *IL6ST* gene. Different colors were used to represent matching cell types. The grey shade represent ±5kb away from rs7731626 SNP in RA GWAS and plots related to T cells.

Additional colocalization results between RA GWAS and OneK1K sc-eQTLs are presented in **Figure S32**. We identified 43 eGene colocalizing with RA (PP.H4 > 0.9), primarily in T and B cells (consistent with literature ^79^). We notably found that *EOMES* eQTLs colocalized with RA in CD8+ T cells (PP.H4 = 0.97), consistent with the role of *EOMES* as a key TF for mediating immunity function in effector CD8+ T cells^80^. Similarly, *CD40* eQTLs colocalized with RA in Plasma cells, reflecting that *CD40* signaling pathway plays an essential role in immune response in B cells ^81,82^.

Altogether, these results illustrate that sc-eQTL analysis can reveal new candidate genes for diseases that are masked by bulk approach.

## Discussion

We developed jaxQTL, an efficient and powerful approach for large-scale eQTL mapping on single-cell data using count-based models. Through simulation and real data analyses, we showed that the negative binomial model was the most powerful and well-calibrated model compared with the linear and Poisson models. In an application to OneK1K, we found that jaxQTL was more powerful in identifying eGenes compared to tensorQTL (linear model) and SAIGE-QTL (Poisson mixed model) while exhibiting comparable or better runtimes when performed using GPUs/TPUs.

We further leveraged jaxQTL results to characterize eGenes and their corresponding fine-mapped sc-eQTLs. First, we found that eGenes are depleted of loss-of-functions variants and large-effect eQTLs, consistent with previous works on bulk-eQTLs ^49,51–53^. Second, we showed that sc-eQTLs are more distal to TSS and more enriched in cell-type matched CREs compared with bulk-eQTLs, and that sc-eQTLs effects are largely consistent across cell types (as observed using eQTLs from bulk cell type samples in ref.^37^), with cell-type-specificity increasing with distance to TSS. These results summarize that while bulk-eQTLs identify primarily proximal regulatory effects with low cell-type-specificity (e.g., promoter), sc-eQTLs allow to identify additional distal regulatory effects with medium to high cell-type-specificity (e.g., enhancer). Finally, we demonstrated that sc-eQTLs explain more heritability than bulk-eQTLs for GWAS traits, suggesting that the GWAS risk variants were partially driven by eQTLs with medium to high cell - type-specificity. We used an example of *ANKRD55-IL6ST* loci to demonstrate that sc-eQTLs can prioritize RA-associated gene *IL6ST* missed by bulk approach. This candidate gene is well-documented for its functional role on the key therapeutic target cytokine *IL6* ^*83–85*^, and is further supported by other genetic evidence such as its higher PoPs prioritization scores ^86^ compared with the closest gene *ANKRD55* (top 1% vs 13% percentile respectively).

jaxQTL has several advantages compared to existing sc-eQTL methods. First, jaxQTL requires no data transformation on gene expression outcomes such that it provides an eQTL effect estimate on the interpretable count data scale. Second, we showed through simulations that jaxQTL outperformed the linear model (used by tensorQTL) in identifying eQTLs, especially for lower expressed genes and rare cell types. This will be well suited for rare cell types and precise cell states in short-read scRNA-seq and low counts of isoforms transcripts in long-read scRNA-seq. Lastly, jaxQTL leverages GPU/TPU to maximize efficiency in sc-eQTL mapping at population scale (N>100) while performing the permutations requested in eQTL best practices ^28^. Although SAIGE-QTL similarly used a count-based model without permutation calibration, jaxQTL on GPU/TPU was on average 3.7x faster than SAIGE-QTL with sample size observed (N=982) in OneK1K and 1,560x faster in simulated data for dominant cell type with sample size N=10,000 (mimicking the upcoming TenK10K data). Importantly, when performing ACAT-V to compute gene-level P values, jaxQTL was 1.3 - 10,596x times faster than SAIGE-QTL for N=100 to 10,000, and displayed times comparable with the linear model of tensorQTL (with permutations).

Our findings have several implications for further single-cell sequencing studies and their integration with GWAS. First, we demonstrated the benefits of the negative binomial model over the Poisson mixed model for sc-eQTLs mapping. jaxQTL can be applied to other count data such as peak reads from scATAC-seq, or extend to accommodate other molecular outcomes such as isoform ratios, while efficiently accounting for the larger number of tests to perform in these datasets (e.g. ∼300K of ATAC-seq peaks in scATAC-seq ^87^ vs. ∼20K genes in scRNA-seq). We thus recommend considering jaxQTL with the negative binomial models for further analyses of population-scaled scATAC-seq and/or multiome datasets to identify chromatin activity QTLs. Second, jaxQTL is scalable for identifying sc-eQTLs in extremely large scRNA-seq datasets. As advances in the ability to multiplex samples in single-cell sequencing assays is allowing generating scRNA-seq and multiome datasets in hundreds to thousands of samples, we expect jaxQTL to be leveraged for analyzing datasets beyond OneK1K and TenK10K. Third, we highlighted that sc-eQTL effects are more shared across cell types than previously reported ^19^. This property might motivate new sc-eQTL mapping and fine-mapping methods jointly integrating all cell types to increase power. Finally, we observed that while OneK1K sc-eQTLs explained a higher proportion of heritability than bulk-eQTLs for GWAS of immune traits, they did not close the missing link between GWAS and eQTLs. While generation of larger scRNA-seq datasets will improve the detection of distal sc-eQTLs with high cell-type-specificity, closing this gap might involve identifying sc-eQTLs with *trans*-effects, generating scRNA-seq data from disease-relevant contexts (such as stimulating condition ^88^ and developmental stage^89^), or generating other types of single-cell QTLs (such as splice QTLs^90^, chromatin activity QTLs^87^ and methylation-QTL ^91^). In all those scenarios, jaxQTL can be easily extended to efficiently analyze those datasets.

We note several limitations of our work. First, jaxQTL power is dependent on cell type abundance, which will limit sc-eQTL power for rare cell types and precise cell states. Despite this, jaxQTL identified more eGenes within rare cell types than existing methods. Second, pseudobulk approaches aggregate read counts over discrete cell types, which may fail to capture dynamic contexts, thus limiting the identification of dynamic sc-eQTLs^34,35^ (i.e. variants impacting gene expression within a cell type whose effects vary dynamically along a continuous state). Identifying dynamic sc-eQTLs with jaxQTL could be performed by identifying sc-eQTLs in more precise cell states, but further work would be required to evaluate this approach. Third, our analyses were restricted to PBMCs and immune-related diseases, and it is unclear if our conclusions translate to sc-eQTLs from different tissues and disease types. However, ongoing release of sc-RNAseq data from brain samples will allow characterizing brain sc-eQTLs and their role in psychiatric traits^20^. Fourth, our analyses were also restricted to European individuals. Assessing the transportability of our conclusions in non-European populations is a critical future research direction, as different environments and genetic backgrounds impact gene regulation and disease effect sizes ^92^. While the recent release of a population-scaled scRNA-seq from East-Asian individuals will allow identifying sc-eQTLs and characterizing their role in East-Asian populations^90^, there is a need to generate datasets in more diverse populations. Despite these limitations, jaxQTL provides an efficient and flexible framework for eQTL mapping on single-cell data using a count-based model. Our findings enable rich biological and mechanistic interpretation for disease risk loci at the cell-type level and nominate therapeutic targets for complex diseases.

## Data Availability

Single-cell eQTL summary statistics results produced by jaxQTL and fine-mapping results are available on Zenodo:10.5281/zenodo.14624945

https://github.com/mancusolab/jaxQTL

https://github.com/mancusolab/jaxqtl_analysis

https://www.encodeproject.org

## Acknowledgments

This work was funded in part by the National Institutes of Health (NIH) under awards P01CA196569, R01HG012133, R01CA258808, R35GM147789, R01GM140287, and R00HG010160. We thank Dr. Joseph Powell for kindly sharing their processed OneK1K gene expression count matrix and genotype data. We thank the ENCODE consortium for generating experimental data for epigenetic markers by cell types. We thank members from the Impact of Genomic Variation on Function (IGVF) consortium QTL focus group for insightful discussions on these results. We thank participants for providing feedback for this work during the Computational Genomics Summer Institute (CGSI) program at UCLA under award GM135043. We thank Dr. Bolun Li for providing user feedback on jaxQTL software.

## Author Contributions

Z.Z., N.M., and S.G. conceived the study. Z.Z., S.G., and N.M. developed the method. Z.Z. performed analyses. N.S. and A.K. prepared data and performed analyses. All authors edited and approved the manuscript.

## Web Resources

JAX: https://github.com/google/jax

tensorQTL: https://github.com/broadinstitute/tensorQTL

SAIGE-QTL: https://github.com/weizhou0/qtl

bedtools: https://bedtools.readthedocs.io/en/latest/

qvalue R package: https://github.com/StoreyLab/qvalue

susieR package: https://github.com/stephenslab/susieR

GTEx pipeline: https://github.com/broadinstitute/gtex-pipeline/tree/master

SLDSC software: https://github.com/bulik/ldsc

1000 Genome annotations: https://alkesgroup.broadinstitute.org/LDSCORE/

PLINK software: https://www.cog-genomics.org/plink/2.0/

mashr: https://stephenslab.github.io/mashr/index.html

GTEx v8 eQTL summary statistics (EUR): https://gtexportal.org/home/downloads/adult-gtex/qtl

DICE cis-eQTL summary statistics: https://dice-database.org/

eQTL catalogue: https://www.ebi.ac.uk/eqtl/

CLUES data: Gene expression data are available in the Human Cell Atlas Data Coordination Platform (https://explore.data.humancellatlas.org/projects/9fc0064b-84ce-40a5-a768-e6eb3d364ee0/project-matrices) and at GEO accession number GSE174188. Genotypes are available at dbGap accession number phs002812.v1.p1

GTEx v8 SuSiE fine-mapping results: https://www.finucanelab.org/data

Gene GTF file (release 84): https://ftp.ensembl.org/pub/grch37/release-84/gtf/homo_sapiens/Homo_sapiens.GRCh37.82.gtf.gzpLI and LOEUF score from gnomad:

https://storage.googleapis.com/gcp-public-data--gnomad/release/4.0/constraint/gnomad.v4.0.constraint_metrics.tsv

## Data and Code Availability

jaxQTL software: https://github.com/mancusolab/jaxQTL

jaxQTL analysis code: https://github.com/mancusolab/jaxqtl_analysis

Original Onek1k data are available at: https://www.ncbi.nlm.nih.gov/geo/query/acc.cgi?acc=GSM5901755

We downloaded the call sets from the ENCODE portal (http://www.encodeproject.org/)with the following identifiers: ENCFF313TWH (CD4 T cell), ENCFF635YOQ (CD8 T cell), ENCFF071MEQ (B cell), ENCFF814VKT (NK cell), and ENCFF468QFA (Monocyte).

## Online Methods

### GLM and count-based eQTL models

Here, we describe the core generalized linear model (GLM) for a focal gene within a focal cell type *c*, assuming that cell type labels for each cell have been provided. Specifically, jaxQTL models the conditional expectation of pseudobulk counts *y*_*c*_ (i.e. sum of read counts across cells within the cell type), given covariates *X*(e.g., age, sex, genotyping principal components) and a cis-genetic variant *g*, as

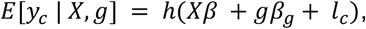

where *β*are the covariate effects, *β*_*g*_ is the allelic effect, *l*_*c*_ is an offset adjusting for differences in library size, and *h*(·) is a function which maps linear predictions to expected values. Additionally, jaxQTL models the conditional variance as,

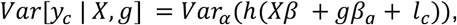

where *Var*_*α*_(·) corresponds to a variance function determined by the specified likelihood (e.g., Poisson, negative binomial) allowing for an overdispersion parameter *α* if required (e.g., negative binomial) ^45,93^.

We fit a null GLM using iteratively reweighted least squares (IRWLS). Namely, assuming there is no genotype effect (i.e. *β*_*g*_ = 0), the update step for covariate effects *β*at the *t* + 1 iteration can be computed by solving a linear system given by,

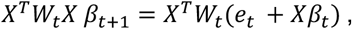

where *e*_*t*_ ∝ (*y* − *μ*_*t*_) are the “working residuals”, *μ*_*t*_ = *h*(*Xβ*_*t*_ + *l*_*c*_), and *W*_*t*_ are the GLM weights proportional to the reciprocal of the conditional variance. To solve this linear system, jaxQTL allows for different solvers (e.g., QR, Cholesky, and conjugate gradient), however in practice we found Cholesky to outperform other approaches.

While both the Wald test and score test are implemented in the software, jaxQTL employs the score test in assessing the nonzero cis-SNP effect *β*_*g*_ on *y*_*c*_ for its improved computational efficiency^94^. Specifically, we first fit the null GLM model described above. Next, jaxQTL uses a block matrix approach to efficiently compute score test statistics for all *p* cis-SNPs in a focal gene. Let 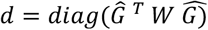 be a length *p* vector, then the vector of test statistics *Z* is given by,

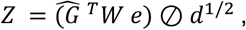

where *Ĝ* is the weighted residualized genotype obtained by *Ĝ* = *G* − *X*(*X*^*T*^*WX*)^−1^*X*^*T*^*WG, d*^1/2^ is the element-wise square-root, and ⊘ is element-wise division.

To adjust for multiple testing corrections in eGene discovery, jaxQTL provides gene-level *P* values calibrated by a permutation-based Beta-approximation approach, which is similarly implemented in FastQTL and tensorQTL software ^28,29^. To infer beta distribution parameters, we applied natural gradient descent using second order approximation to ensure parameters stay on the manifold (i.e. > 0)^95^. Lastly, we controlled the false discovery rate (FDR) and identified eGenes using these gene-level *P* values.

We implemented jaxQTL in Python to enable *just-in-time* (JIT) compilation through the *JAX* package (**Web Resources**), which generates and compiles heavily optimized C++ code in real-time and operates seamlessly on CPU, GPU, or TPU (**Code Availability**).

### Analysis of OneK1K single-cell data

We obtained 1,267,768 PBMC blood cells for 14 cell types from *N* = 982 healthy individuals of European ancestry in the OneK1K cohort^19^. Each donor has an average of 1,291 cells (range 62-3,501). Each cell type has a varying number of donors due to sampling variance (**Table S2**). For sc-eQTL mapping, we created pseudobulk count data for each of the pre-annotated 14 cell types. After retaining genes with sample-coverage (i.e., fraction of non-zero expression read counts) in at least 1% of the population for each cell type, we performed sc-eQTL mapping for an average of 16,096 genes per cell type (**Table S2**). To establish a baseline for comparison, we created “bulk” data by summing all read counts across cell types for every gene and identified bulk-eQTLs using jaxQTL.

We first aimed to benchmark between different sc-eQTL models, including linear, negbinom, and Poisson. For negbinom and Poisson, we calculated individual library size in each cell type, i.e., the offset term in the model, by summing read counts across all genes per individual. For the linear model, we normalized gene expression read counts between individuals by TMM approach and then normalized across individuals by rank-based inverse normal transformation, as performed in GTEx ^10^. We implemented all three models using a score test in jaxQTL.

For genotype data, we retained 5,313,813 SNPs with imputation INFO score >0.8, MAF>0.05, and Hardy Weinberg equilibrium (HWE) *P*>1e-6. Genotype PCs were calculated using 459,603 LD-pruned SNPs with INFO>0.9, MAF>0.01, and HWE *P*>1e-6. Across all sc-eQTL models, we adjusted covariates including age, sex, first six genotype PCs, and two expression PCs computed in each cell type. We defined the cis-window size as ±500kb (total 1Mb) around TSS. We downloaded the gene annotation GTF file (Homo_sapiens.GRCh37.82) and collapsed it to a single transcript model using “collapse_annotation.py” from GTEx analysis pipeline (**Web Resources**)^10^. We controlled gene-level FDR at 0.05 per cell type using the qvalue method on gene-level *P* values through *qvalue* R package (**Web Resources**) ^96^. eGenes were identified by qvalue < 0.05. We used genome build hg19 for all variants and gene annotations.

To benchmark with other existing software, we compared the eGenes results of jaxQTL against tensorQTL ^29^ and SAIGE-QTL ^36^. We first restricted genes to sample-coverage > 10% as in ref. ^36^ and obtained their SAIGE-QTL eGenes results ^36^. Then we applied FDR control on this subset of genes to call eGenes for jaxQTL-negbinom. Similarly, we performed sc-eQTL mapping using tensorQTL on this set of genes (**Web Resources**). We note that the tensorQTL results we report are different from tensorQTL results reported in ref. ^36^, as we sum pseudobulk counts (following GTEx recommended guidelines to transform counts from bulk RNA-seq ^10^) rather than averaging them. All reported P values are two-sided unless specified otherwise.

### Simulations

To evaluate the performance of the jaxQTL-linear, jaxQTL-negbinom, jaxQTL-Poisson, tensorQTL (linear), and SAIGE-QTL (Poisson mixed effect) models, we first simulated read counts *y*_*ij*_ for individual *i* in cell *j* for a focal gene under the Poisson mixed effect model, given by:

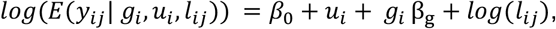

where *β*_0_ is a baseline intercept, *u*_*i*_ *∼ N*(0, *σ*^2^_*u*_) is the random intercept for individual *i* that induces within-sample correlation across cells, *g*_*i*_ is genotype with effect-size *β*_*g*_ *∼ N*(0, *h*^2^_*cis*_) where *h*^2^_*cis*_is cis-SNP heritability. To reflect the cell-wise and individual-wise read counts (i.e., library size *l*_*ij*_) observed in scRNA-seq data, we sampled *l*_*ij*_ from empirical values observed in OneK1K data. To fit pseudobulk linear, negbinom, and Poisson models, we created pseudobulk counts as *y*_*i*_ = ∑_*j*_ *y*_*ij*_ and library size as *l*_*i*_ = ∑_*j*_ *l*_*ij*_. We varied the baseline expression *β*_0_, cis-SNP heritability *h*^2^_*cis*_, the random intercept variance *σ*^2^_*u*_, MAF, and sample size N. Given fixed *β*_0_ values, we obtained varying sample-coverage across simulation replicates and calculated the average simulated sample-coverage (**Figure 1**; **Figure S4-8**).

To evaluate the performance under model misspecification, we simulated single-cell read counts from the standard Poisson model assuming no within-individual correlation between cells, i.e., *σ*^2^_*u*_= 0. In each scenario, we performed a score test for association between simulated gene expression and genotype after fitting jaxQTL-linear, jaxQTL-Poisson, jaxQTL-negbinom, and SAIGE-QTL models. Different from the score test in jaxQTL-linear, tensorQTL reports Wald test statistics. For the linear models (jaxQTL-linear and tensorQTL), we normalized pseudobulk counts by rank-based inverse normal transformation ^10,97^. Each simulation had 500 replicates.

### Replication of sc-eQTLs

To validate sc-eQTLs identified by jaxQTL-negbinom, we performed replication analysis using jaxQTL on two independent cohorts from CLUES study ^47^. We obtained scRNA-seq for N=256 individuals of European and Asian ancestry (see **Web resources**). We removed 50 control individuals from the ImmVar study, 2 outliers detected through a PCA analysis and 2 male individuals from the remaining based on ref ^92^. After intersecting with genotype data, we retained 88 European- and 88 Asian-ancestry individuals for replication analysis. Of these, 65 and 67 individuals were diagnosed with systemic lupus erythematosus (SLE) but were not in active state of disease flare. We matched 7 cell types in CLUES with 14 cell types in OneK1K. We performed analysis in European and Asian individuals separately. For lead SNP-eGene pairs identified by jaxQTL-negbinom, we fitted negbinom model using jaxQTL in CLUES cell types. We adjusted for age, sex, first six genotype PCs, SLE status, and batch numbers in sc-eQTL model. For each cell type, we reported the fraction of pairs replicated at FDR < 0.05 using *qvalue* R package^96^. To compare sc-eQTL effect size estimated by jaxQTL in CLUES and OneK1K samples adjusting for fitted count scale differences, we calculated an adjusted slope estimate as 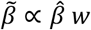 where 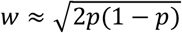 after accounting for weights in the GLM. Lastly, we compared results reported in the original OneK1K linear-model based analyses to demonstrate directional consistency in sc-eQTL effect estimates^19^.

We further investigated the replication of lead SNP-Gene pairs in eQTLs identified by previous bulk-eQTL and sc-eQTL studies (see **Web resources**). For bulk-eQTL studies, we downloaded 1) cis-eQTL results from European whole blood samples in GTEx v8 (N=588)^10^; 2) cis-eQTL summary statistics results from FACS-sorted PBMC immune cell types in DICE study (N=91) ^48^. For sc-eQTL study, we downloaded cis-eQTL summary statistics from PBMC scRNA-seq data in CLUES study curated by eQTL catalogue (N=193) ^47,98^. All these prior results are based on the linear model approach when performing eQTL mapping. Again we reported the fraction of pairs replicated at FDR < 0.05 within each cell type.

### Computational runtime

To evaluate the computational runtime of jaxQTL on cis-eQTL mapping in comparison with other software, we randomly selected 50 genes from chromosome 1 with sample-coverage > 1% in CD4_NC_, B_IN_, and Plasma cells observed in OneK1K. To benchmark the performance across different sc-eQTL sample sizes, we performed downsampling to create gene expressions for N=100, 300, 500, and 700 individuals. Moreover, we performed upsampling for the expected TenK10K cohort^21^. Specifically, we sampled N=10,000 individuals using the single-cell data matrix. Assuming each individual has a total of 5,000 cells as expected in TenK10K, we sampled the number of cells per person proportionally as observed for these three cell types. Then we created pseudobulk data for jaxQTL and tensorQTL.

### Fine-mapping on sc-eQTLs

To identify causal eQTL for every eGene, we performed fine-mapping using SuSiE summary statistics approach for eGenes identified by sc-eQTL and bulk-eQTL approach (both jaxQTL-negbinom). We excluded eGenes in the *MHC* region (chr6: 25Mb-34Mb) with complex LD patterns and the *MAPT* region (chr17: q21.31) with complex inversion and duplication ^99,100^. For optimal statistical power, we first used jaxQTL-negbinom to compute all pairwise summary statistics for cis-SNPs in every eGene. We calculated the in-sample LD correlation matrix for cis-SNPs 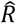 after projecting out the covariates effect under the GLM weights. Specifically for negbinom model results, we calculated a weighted residualized *G* by *Ĝ* = *G* − *X*(*X*^*T*^*WX*)^−1^*X*^*T*^*WG*, followed by computing 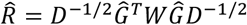, where *D* = *diag*(*Ĝ*^*T*^*WĜ*) and *W* is the individual weights calculated after fitting the null model.

### Enrichment analysis on sc-eQTLs

We downloaded annotations from the LDSC baseline model and selected 12 annotations of promoter-like regions, enhancers, conserved regions, and epigenetic markers (**Web Resources**). For cell-type matched candidate cis-regulatory elements (CREs), we downloaded CRE peaks in PBMC cells identified by scATAC-seq^101^. We extracted CREs from cell types matched with cell types in OneK1K based on labels and marker genes (**Table S8**). For enhancers and promoters, we collected 33 samples for 6 cell types from EpiMap and used *bedtools* to merge peak regions from different samples in the same cell type (**Table S9**). Lastly, to interrogate the accuracy of sc-eQTL linked to target genes, we obtained the enhancer-gene links identified by SCENT^57^ in B cells, T/NK cells, and Myeloid cells. Cell types were matched based on labels (**Table S10**). We removed ENCODE promoter-like regions from SCENT peaks to retain putative enhancer regions.

For enrichment analysis on scATAC-seq, EpiMap enhancers, and promoters, we created annotations for cis-SNPs taking the value of 1 if falling within the CRE region and 0 otherwise. For every eGene, we performed logistic regression similar to torus^102^ by fitting *logit*(*PIP*_*k,j*_) = *β*_0_ + *β*_*k, a*_*a*_*k,j*_, where *j* denotes cis-SNPs in eGene *k* and their annotation *a*. To obtain a single enrichment score for every annotation in a cell type, we performed a fixed effect meta-analysis using fitted slopes and their standard errors across all eGenes. Specifically, the meta-analyzed slope over eGenes is (∑_*k*_ *β*_*k,a*_ /*W*_*k,a*_)/(∑_*k*_ *W*_*k,a*_) with variance 1/ ∑_*k*_ *W*_*k,a*_, where *W*_*k,a*_ = 1/*SE*(*β*_*k,a*_)^2^. When comparing sc-eQTLs against bulk-eQTLs enrichment, we meta-analyzed summary statistics across cell types.

To calculate the enrichment of sc-eQTLs in enhancer-gene pairs identified by SCENT, we defined the enrichment score for every eGene as:

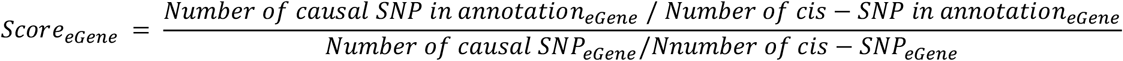

Then we calculated the enrichment for each cell type by taking an average of *Score*_*eGene*_. To evaluate the uncertainty of this mean enrichment score, we calculated the variance of this mean enrichment by bootstrapping 1,000 iterations.

### Cell type sharing of sc-eQTLs

To investigate cell type specificity or sharing of sc-eQTLs, we performed the *mashr* analysis on 2,256 fine-mapped sc-eQTL (PIP≥0.5) with complete summary statistics across 14 cell types ^58^ (**Web Resources**). We first constructed a “finemap sc-eQTL” Z score matrix of size 2,256 × 14. To estimate residual covariance between cell types due to sample overlap, we constructed a null sc-eQTL matrix (21,542 × 14) by randomly sampling from 2 SNPs in every gene with max |Z| < 2 across all cell types.

Following the instructions described elsewhere ^58^, we used a data-driven approach to estimate the covariance matrix of the sc-eQTL effect. In brief, we first used the “finemap sc-eQTL” matrix to create 27 candidate covariance matrices including empirical covariance of Z score, 5 rank-1 approximation to the covariance matrix, rank-5 approximation, and 19 canonical covariance matrices created by *cov_canonical()*. Then we applied *cov_ed()* to estimate the covariance pattern by extreme deconvolution. Lastly, we estimated the mixture weight by fitting the *mashr* model on the null sc-eQTL matrix using 27 covariance patterns and residual covariance. Lastly, we fitted *mashr* on the “finemap sc-eQTL” matrix to obtain posterior effect estimates and local false sign rate (LFSR) using the mixture estimates from above. Since we used the Z score model of *mashr*, we converted the posterior estimate back to the effect scale by multiplying their standard errors as described elsewhere ^58^.

To count for sc-eQTL sharing, we first selected 2,012 sc-eQTLs with LFSR < 0.05, which was similar to FDR control. For each significant sc-eQTL, we called the cell type with the strongest *mashr* effect size as discovery cell type. We considered two types of eQTL sharing: 1) “share by sign” means the other cell type shared the sign of effect with the discovery cell type; 2) “share by magnitude” means conditioning on “share by sign”, the magnitude was within a factor of 2 compared to the discovery cell type.

For enrichment analysis of scATAC-seq peaks, we first used *bedtools subtract -A* recursively to identify peaks exclusive to each cell type, i.e., cell-type-specific peaks. Then we calculated the enrichment score using:

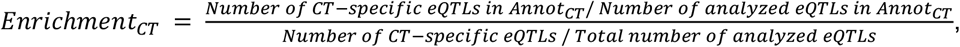

where CT refers to cell type and Annot_CT_ is cell-type-specific peaks. To obtain standard errors for the enrichment, we performed bootstrapping with 1,000 iterations on the cell type labels for CT-specific eQTLs.

### Integration sc-eQTLs with GWASs

To assess the overlap between sc-eQTL and GWAS risk variants, we performed an S-LDSC analysis on 16 GWAS results for blood and immune-related traits (**Web resources; Table S7**). Firstly, we created annotations using SNPs in credible sets of fine-mapped eGenes from 14 cell types (as recommended in ref. ^91^). We constructed three sets of annotations for 1) cell-type sc-eQTL: a union of credible sets of eGenes per cell type; 2) sc-eQTL_union: a union of credible sets from 1) across all cell types; 3) bulk-eQTL: credible sets of eGenes in bulk-eQTL results. Then we annotated SNPs using European individuals from 1000 Genome Project and performed S-LDSC analysis using these annotations (**Web resources**). To estimate heritability, we used baseline-LD v2.2 model (96 annotations) as recommended in ref.^103^ to obtain estimates reducing biases from MAF- and LD-dependent architectures. To identify the likely causal cell types associated with each GWAS trait, we fitted the baseline model v1.2 (53 annotations) to optimize statistical power as recommended in ref.^104^. To account for background non-cell-type-specific eQTLs in the baseline model, we construct an additional annotation by taking a union of fine-mapped SNPs in 95% credible sets from SuSiE results across 49 GTEx tissues (**Web resources**) and sc-eQTL_union from our OneK1K results in order to identify cell-type-specific effects. We focused on two metrics: 1) the proportion of heritability explained by each annotation *h*^2^(*C*) from the baseline-LD v2.2 model result, and 2) the standardized coefficient *τ*_*c*_ ^*\**^calculated by:

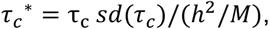

where *τ*_*c*_, *sd*(*τ*_*c*_), *h*^2^ were estimated from the baseline model and *M* was the total number of SNPs that *h*^2^_*g*_ was computed on (*M* = 5,961,159) using 1000 Genome Project and baseline v1.2 model. Here *τ*^*\**^is the change in per-SNP heritability with one standard deviation increase in annotation, which makes it comparable between annotations and GWAS traits. The P values are one-sided hypothesis test for *τ*^*\**^> 0.

