## supplemental figures for "Efficient count-based models improve power and robustness for large-scale single-cell eQTL mapping"

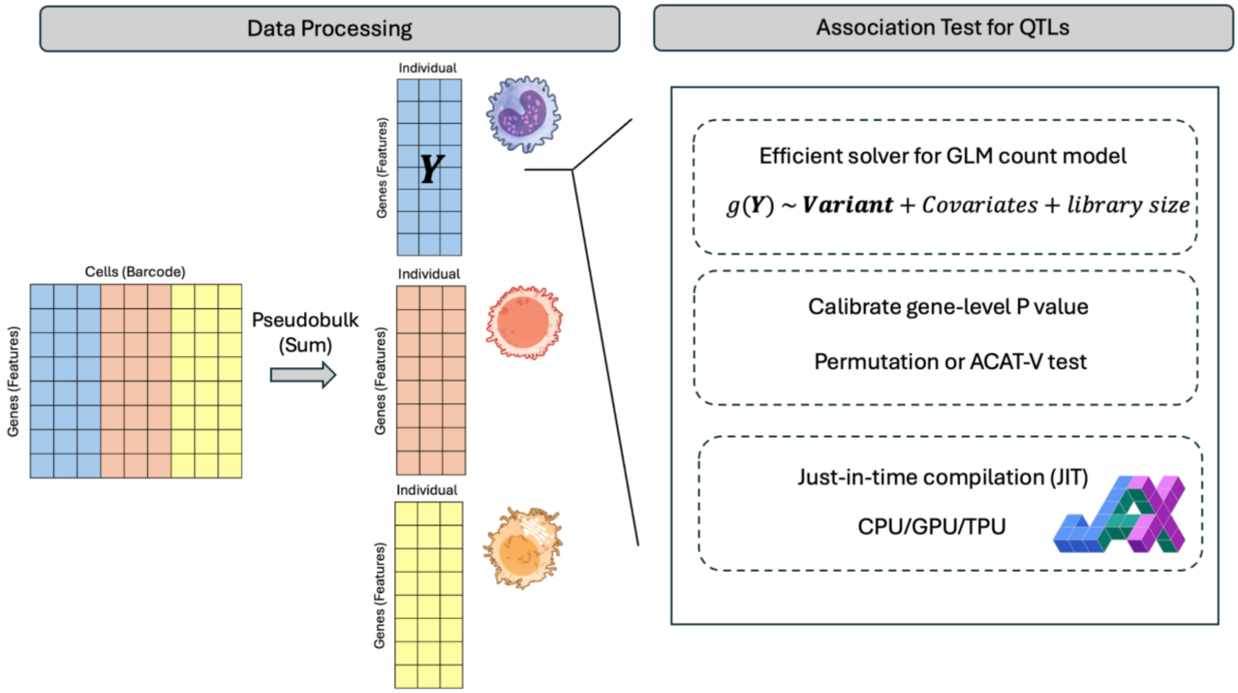


**Figure S1. Overview of jaxQTL.**
jaxQTL requires pseudo-bulking by sum on a single-cell data matrix as input for each pre-annotated cell type. Providing genotype and covariate data for each cell type, jaxQTL can fit a GLM count-based model such as a negative binomial model between gene expression and genotype to identify the lead SNP for each gene (see **Methods**). To compute gene-level *P* value, jaxQTL provides two options: 1) Beta-approximation approach using permutations (recommended), 2) ACAT-V test without permutations (faster). jaxQTL leveraged *just-in-time* (JIT) provided by the JAX python package (**Web Resources**), which is cluster-friendly and operates seamlessly on CPU, GPU, or TPU.

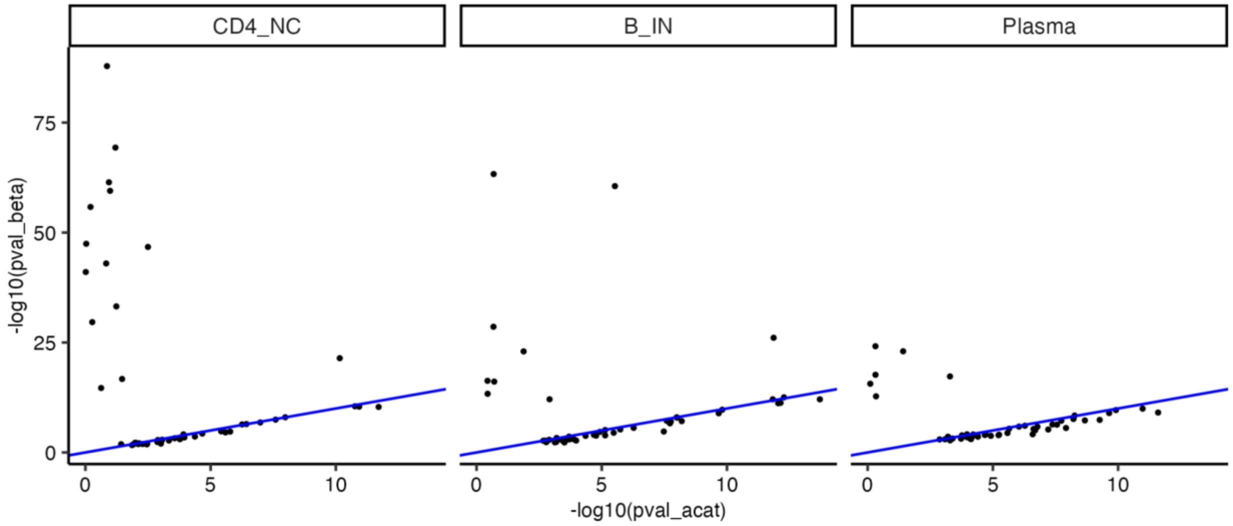


**Figure S2. Permutation-based Beta-approximation method for the negbinom model is more powerful in calibrating gene-level P values than the ACAT-V approach.**
For 50 randomly selected eGenes identified by jaxQTL-negbinom in 3 representative cell types (CD4_NC, B_IN, Plasma), we obtained their gene-level P values using the permutation-based Beta-approximation method (y-axis) and ACAT-V method (x-axis). The identity line is colored in blue.

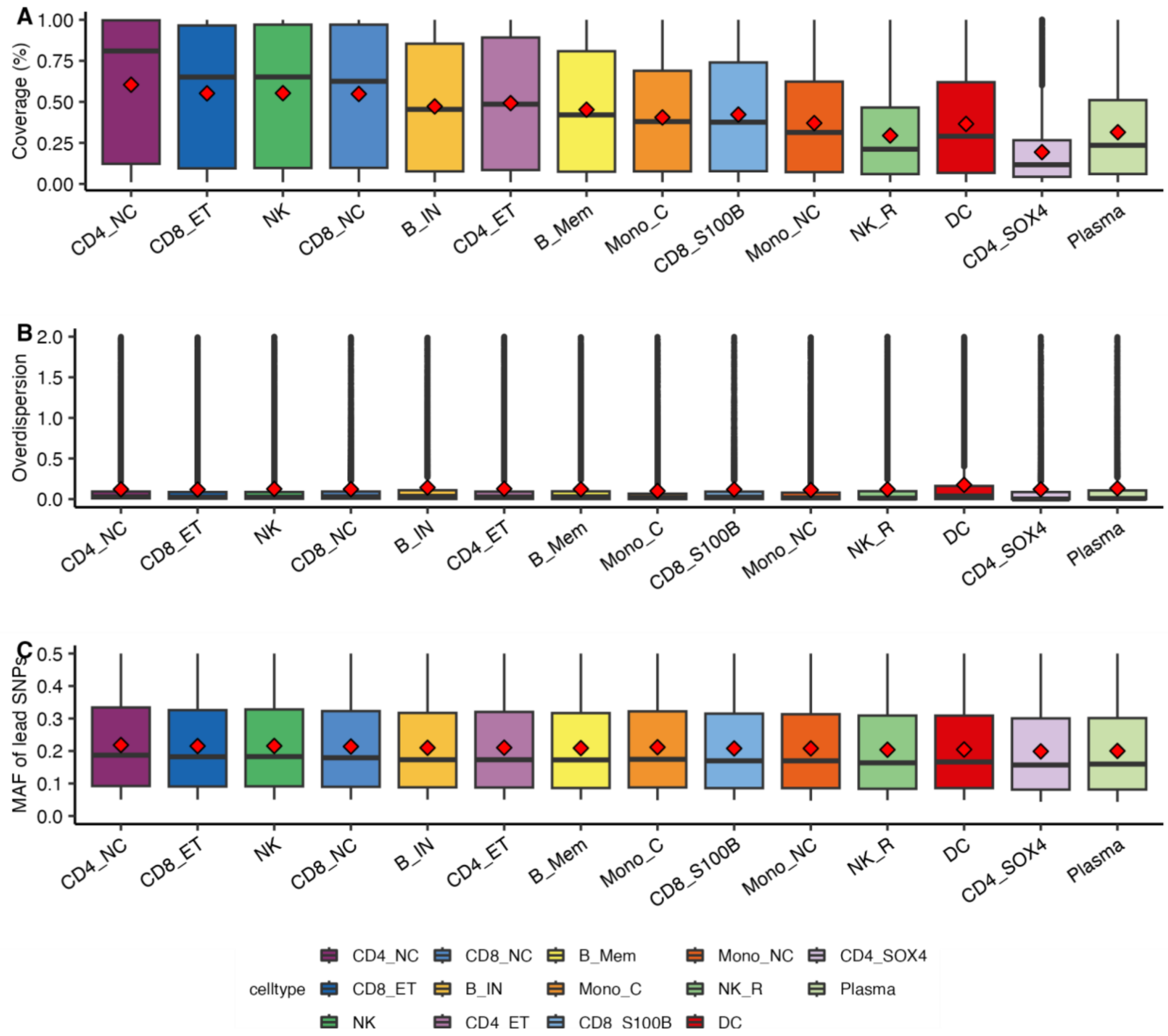


**Figure S3. Distribution of the sample-coverage, estimated overdispersion parameter, and MAF for their lead eQTLs in OneK1K.**

**(A)** We report the sample-coverage (percentage of non-zero expression read counts) of all genes tested for each cell type. **(B)** We report the dispersion parameter estimated in all genes tested from each cell type. Values are truncated at 2.0, with a range from 1e-8 to 90.04. **(C)** We report the minor allele frequency (MAF) of lead SNPs of all genes tested from each cell type. The red diamond is the mean for each boxplot.

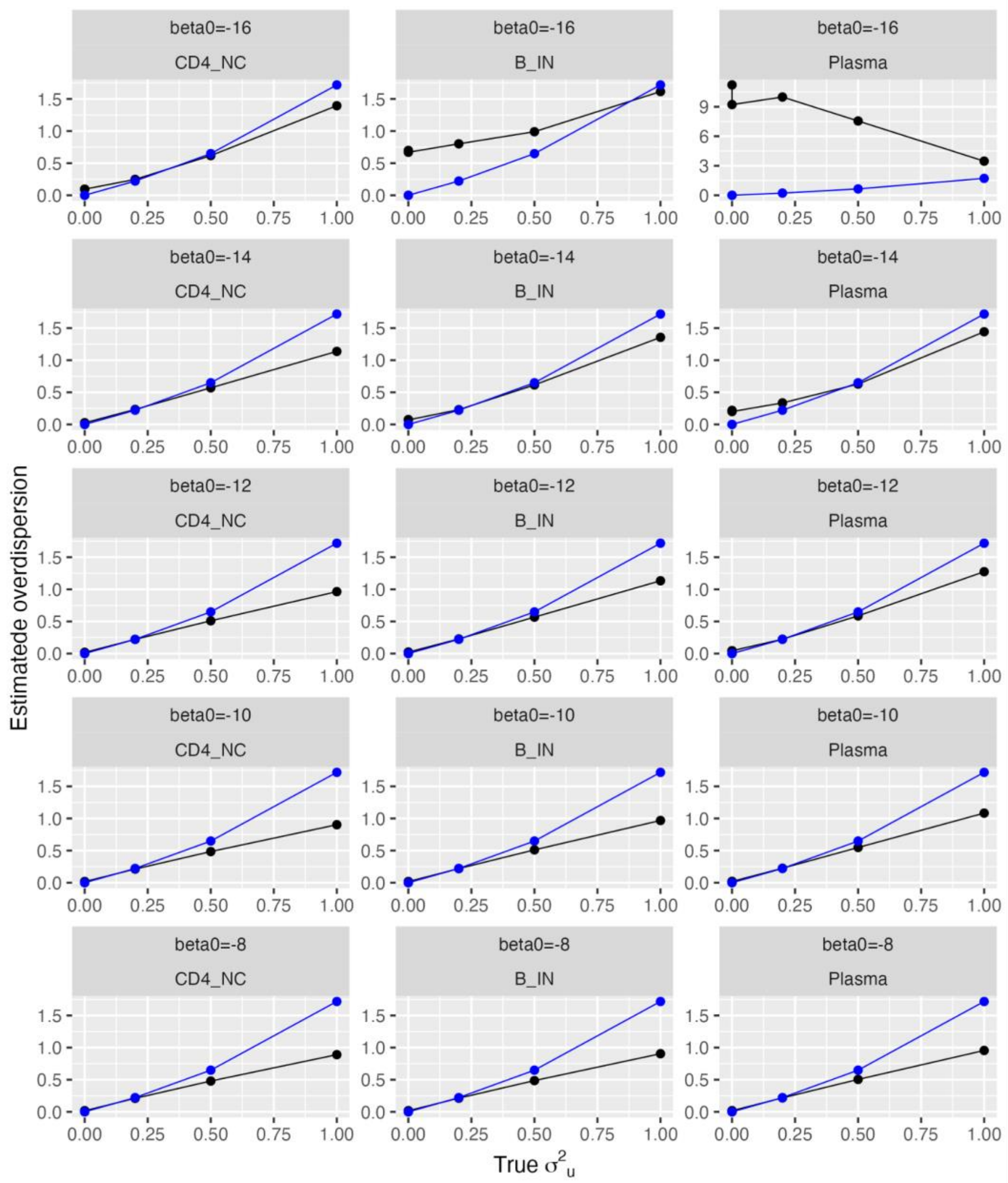


**Figure S4. Estimated overdispersion tracks with the true random intercept variance in the single-cell Poisson mixed effect (PME) model.**

For fixed parameter $h^2_{cis}$= 0.05, MAF = 0.2, and sample size N = 1,000, we generated single-cell data from the Poisson mixed effect model across grid values of true random intercept variance $\sigma^2{}_u$ and true baseline intercept beta0. We plot the estimated overdispersion by pseudobulk jaxQTL-negbinom model and true random intercept variance $\sigma^2{}_u$. We chose $\sigma^2{}_u$= 0.2 to reflect the mean overdispersion observed in OneK1K data (see **Figure S3A**). The blue points are theoretical overdispersion values $exp(\sigma^2{}_u) - 1$ given the structural similarity in variance between negbinom and PME (see **Supplemental Note**).

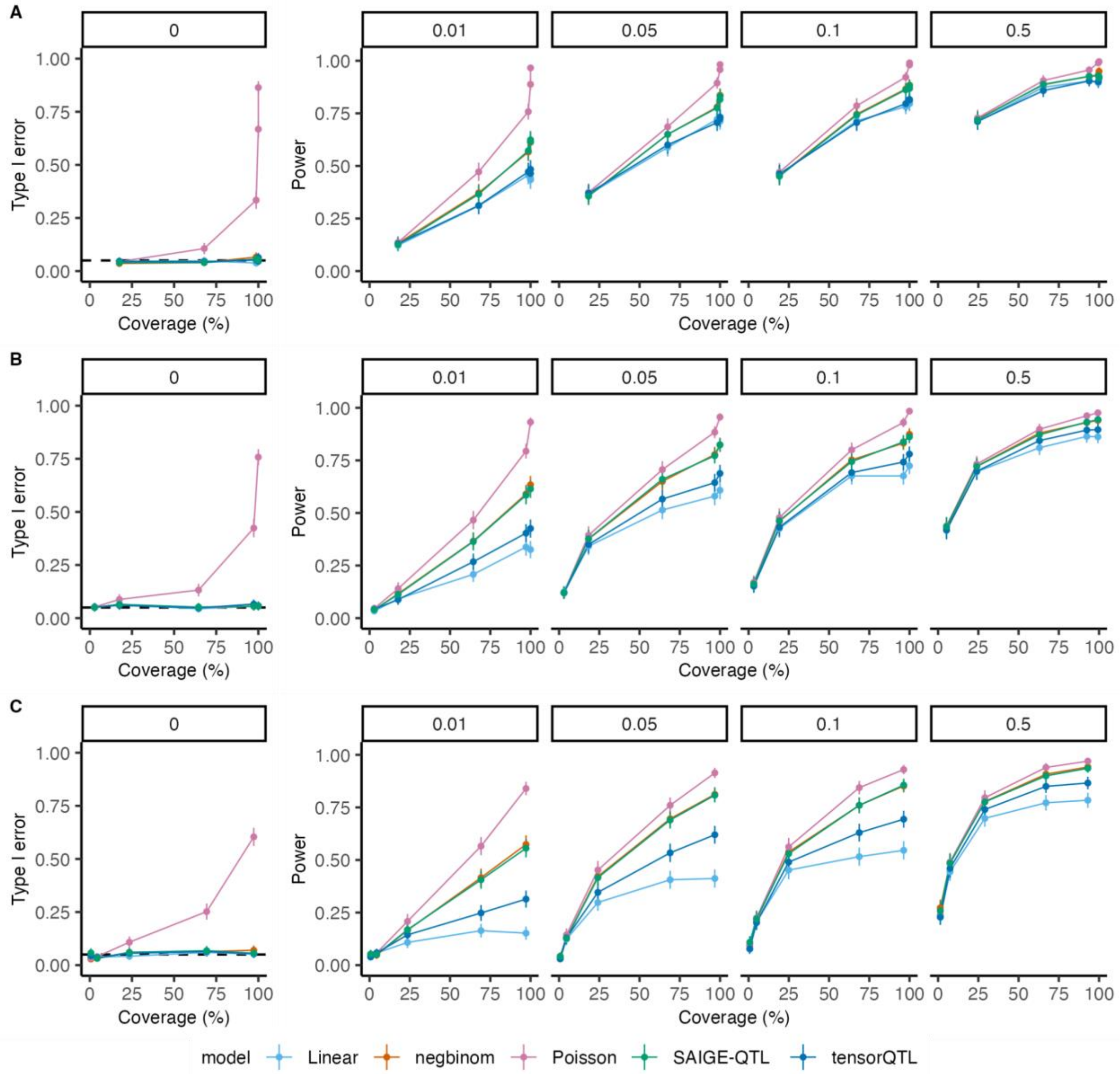


**Figure S5. Simulation results when varying cis-heritability $h^2{}_{cis}$ and sample-coverage.** We report the type I error ($h^2{}_{cis} = 0$) and power for simulations where we varied $h^2{}_{cis}$ (panels) and simulated sample-coverage (x-axis) using individual library sizes sampled from CD4_NC **(A)**, B_IN **(B)**, and Plasma **(C)** cells. We fixed random intercept variance $\sigma^2{}_u$ = 0.2, MAF = 0.2, and sample size N = 1,000. The dashed line on the first column of panels ($h^2{}_{cis} = 0$) is 0.05. Error bars are 95% CIs estimated from 500 replicates.

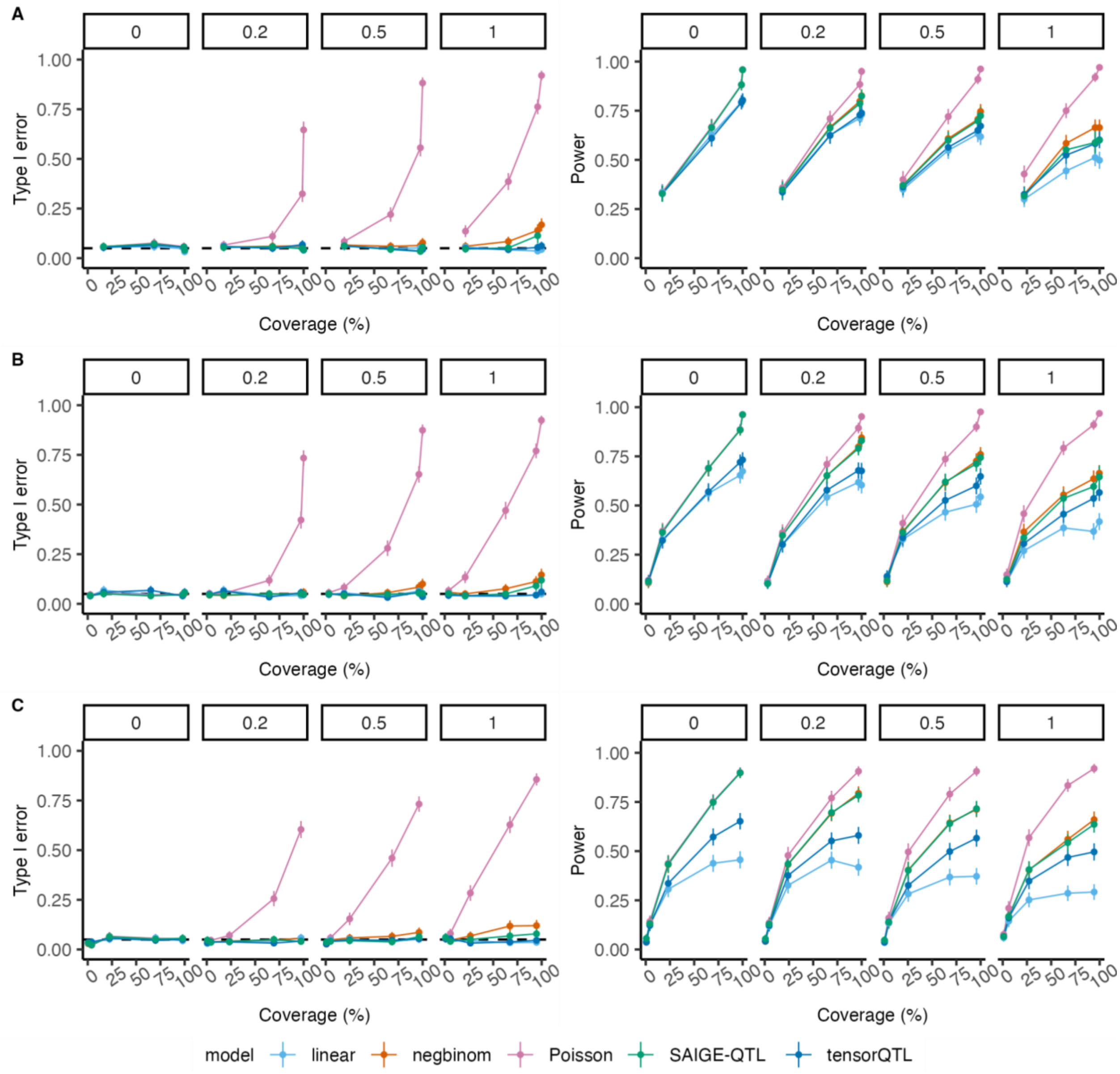


**Figure S6. Simulation results when varying random intercept variance $\sigma^2{}_u$ and sample-coverage.**

We report the type I error ($h^2{}_{cis} = 0$) and power for simulations where we varied $\boldsymbol{V_{re}}$ (panels) and simulated sample-coverage (x-axis) using individual library sizes sampled from CD4_NC **(A)**, B_IN **(B)**, and Plasma **(C)** cells. We fixed $h^2{}_{cis}$ = 0.05, MAF = 0.2, and sample size N = 1,000 for power analysis. The dashed line on the first column of panels ($h^2{}_{cis} = 0$) is 0.05. Error bars are 95% CIs estimated from 500 replicates.

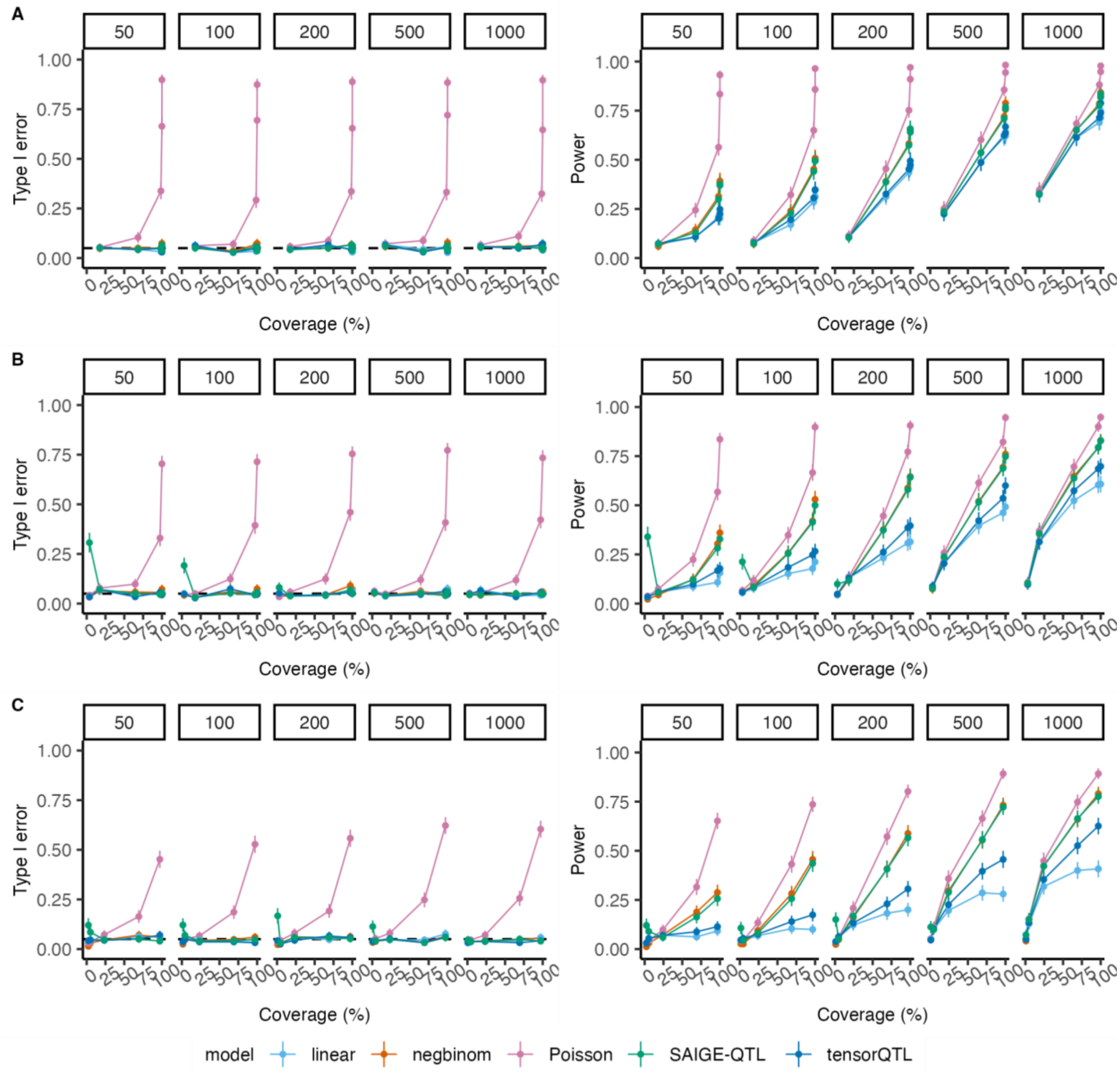


**Figure S7. Simulation results when varying sample size N and sample-coverage.**
We report the type I error ($h^2{}_{cis} = 0$) and power for simulations where we varied sample size N (panels) and simulated sample-coverage (x-axis) using individual library sizes sampled from CD4_NC **(A)**, B_IN **(B)**, and Plasma **(C)** cells. We fixed $h^2{}_{cis}$ = 0.05, random intercept variance $\sigma^2{}_u$ = 0.2, and MAF = 0.2 for power analysis. The dashed line on the first column of panels ($h^2{}_{cis} = 0$) is 0.05. Error bars are 95% CIs estimated from 500 replicates.

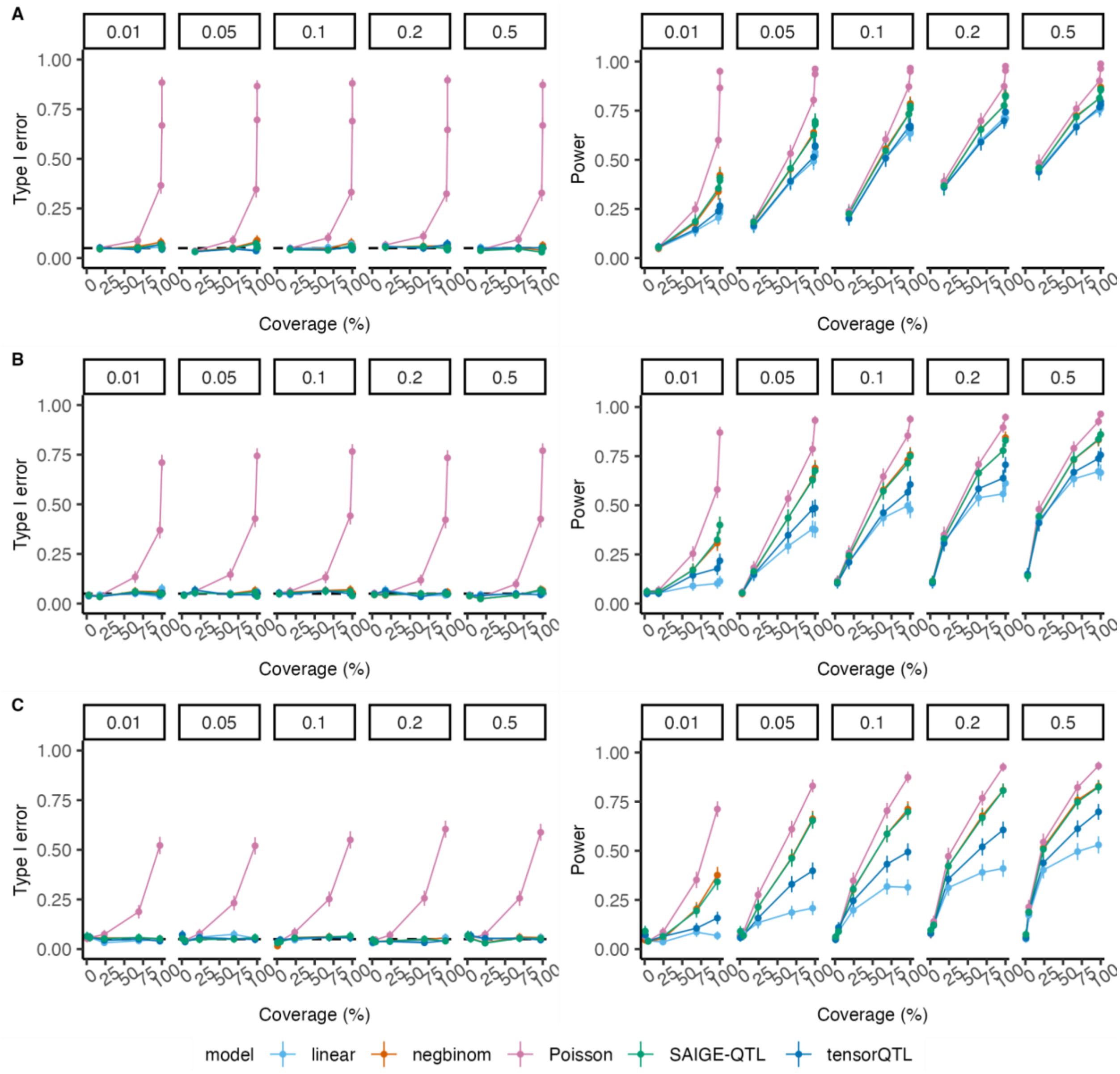


**Figure S8. Simulation results when varying MAF and sample-coverage.**

We report the type I error ($h^2{}_{cis} = 0$) and power for simulations where we varied MAF (panels) and simulated sample-coverage (x-axis) using individual library sizes sampled from CD4_NC **(A)**, B_IN **(B)**, and Plasma **(C)** cells. We fixed $h^2{}_{cis}$ = 0.05, random intercept variance $\sigma^2{}_u$ = 0.2, and sample size N = 1,000 for power analysis. The dashed line on the first column of panels ($h^2{}_{cis} = 0$) is 0.05. Error bars are 95% CIs estimated from 500 replicates.

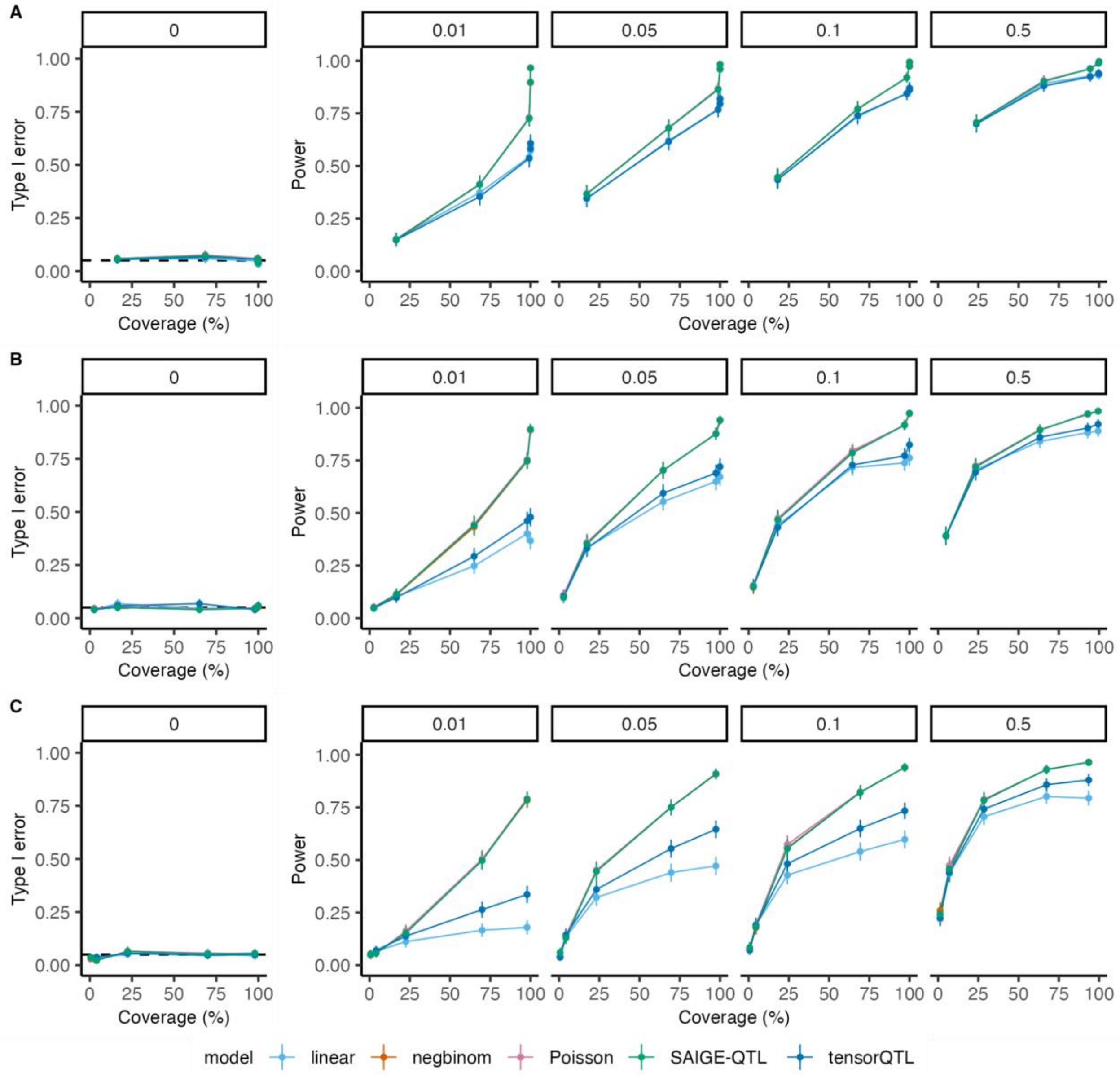


**Figure S9. Simulation results when varying cis-heritability $h^2{}_{cis}$ and sample-coverage under the PME model without random intercept (standard Poisson).**

We report the type I error ($h^2{}_{cis} = 0$) and power for simulations where we varied $h^2{}_{cis}$ (panels) and simulated sample-coverage (x-axis) using individual library sizes sampled from CD4_NC **(A)**, B_IN **(B)**, and Plasma **(C)** cells. We fixed random intercept variance $\sigma^{\mathbf{2}}{}_{\boldsymbol{u}}$ = 0 (Poisson), MAF = 0.2, and sample size N = 1,000. The dashed line on the first column of panels ($h^2{}_{cis} = 0$) is 0.05. Error bars are 95% CIs estimated from 500 replicates.

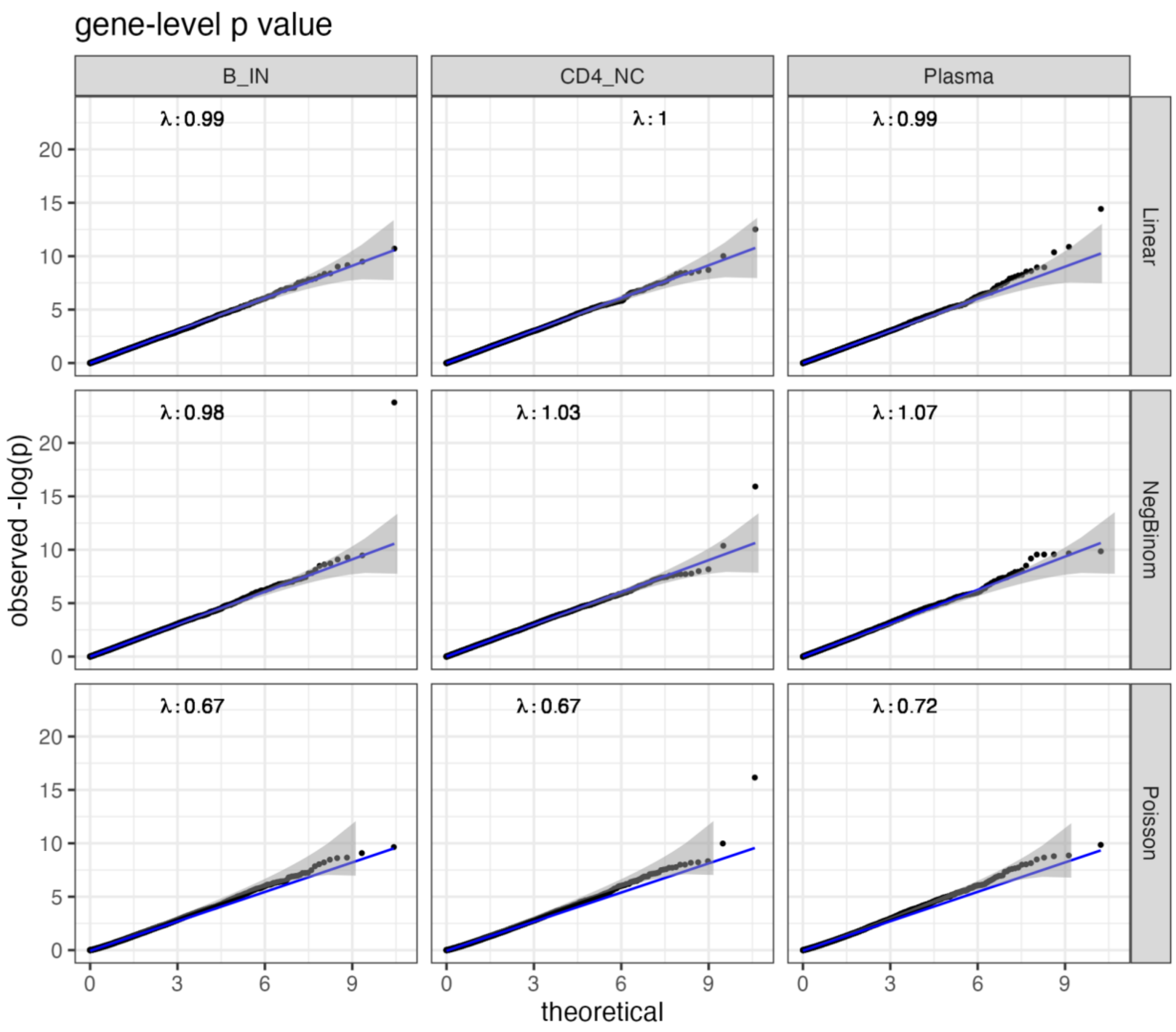


**Figure S10. Gene-level *P* values obtained by the permutation-based approach are well-calibrated in real data.**

We permuted the gene expression for all genes with sample-coverage > 1% in B_IN, CD4_NC, and Plasma cells of OneK1K data and performed cis-eQTL mapping for all genes using linear, negbinom, and Poisson models. We report the QQ plot of these gene-level *P* values and the corresponding genomic inflation factor $\lambda$. Shaded grey is a 95% confidence band. The identity line is in blue.

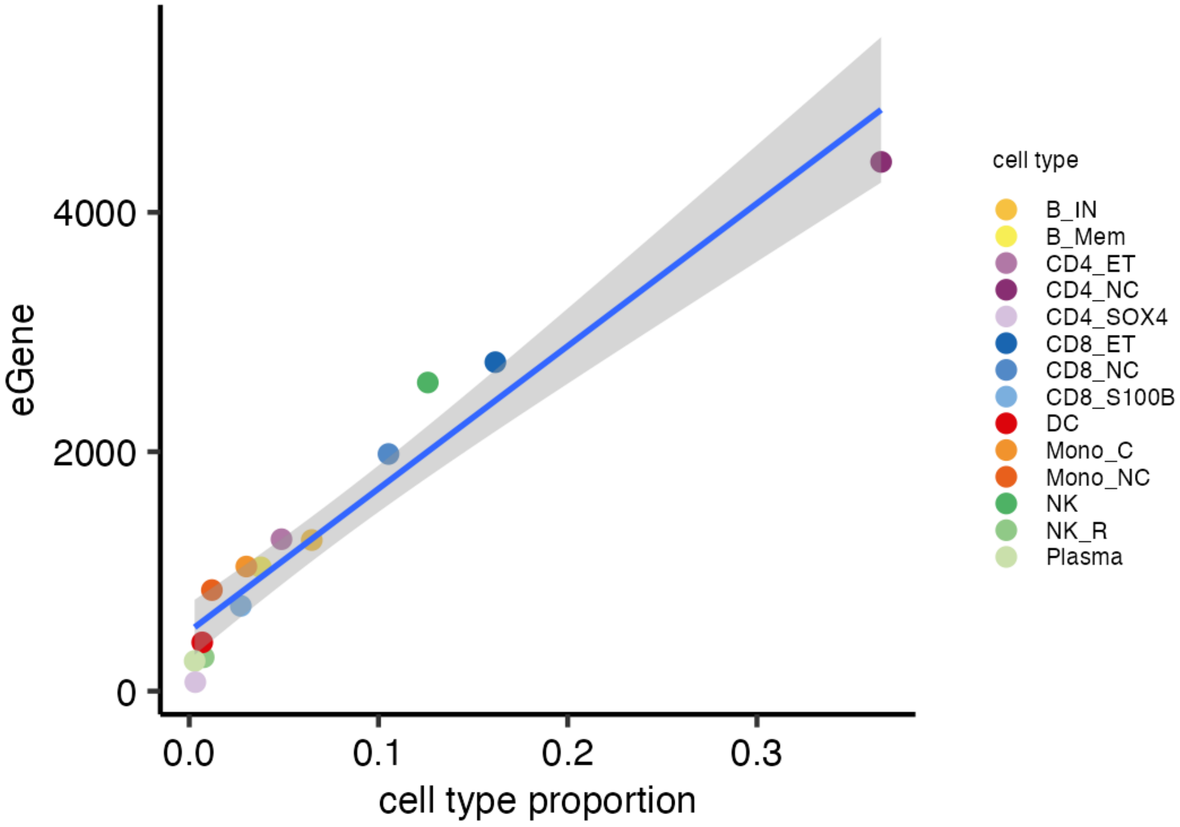


**Figure S11. The number of eGenes identified per cell type tracks with cell type proportions.**
We report the number of eGenes identified by jaxQTL-negbinom per cell type as a function of the cell type proportion. The fitted line is a linear regression with a 95% confidence interval.

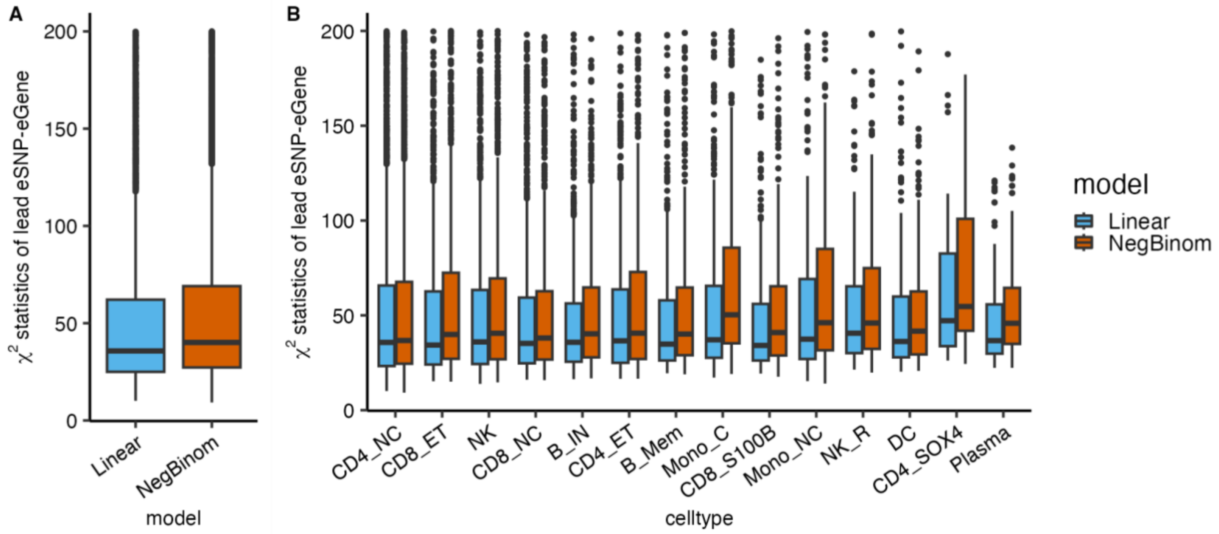


**Figure S12. The negbinom model shows higher $\chi^2$ test statistics than the linear model for lead SNP-eGenes identified by both models.**

For eGenes identified by both jaxQTL-negbinom and jaxQTL-linear, we report the distribution of $\chi^2$ test statistics for all lead SNP-eGenes **(A)** and all cell types **(B)**. y-axes are truncated at 200 for visualization purposes.

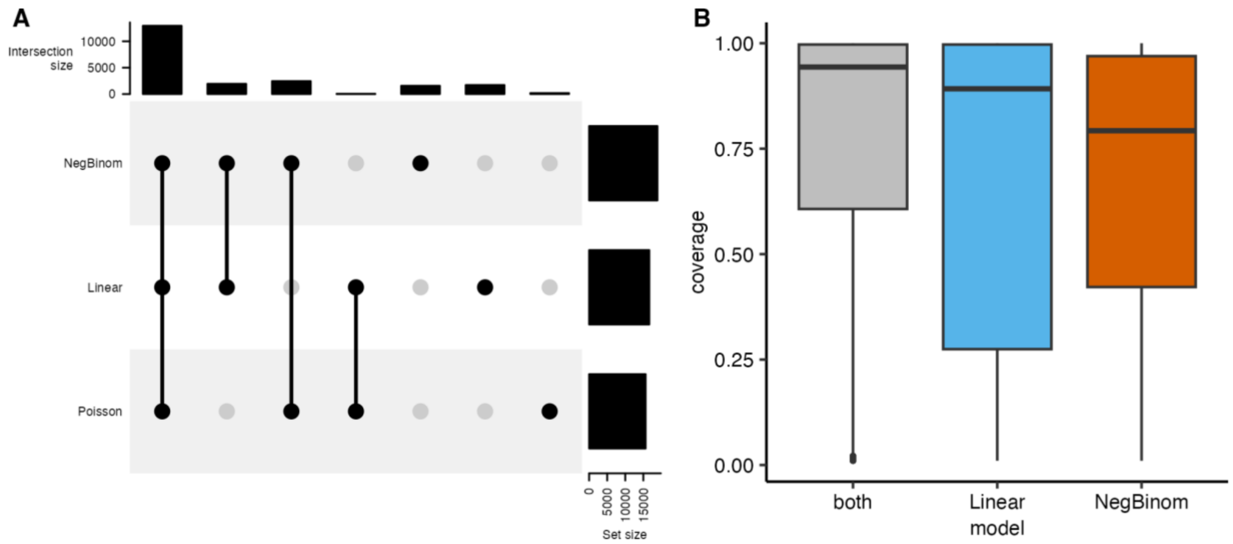


**Figure S13. eGenes that are specific to jaxQTL-negbinom are driven by lower expressed genes.**
**(A)** We report the substantial overlap of eGenes identified between three models in an upset plot. **(B)** Focusing on jaxQTL-negbinom and jaxQTL-linear, we report the sample-coverage (i.e., percentage of non-zero expression read counts) for eGenes identified by either both or specific to jaxQTL-negbinom or jaxQTL-linear.

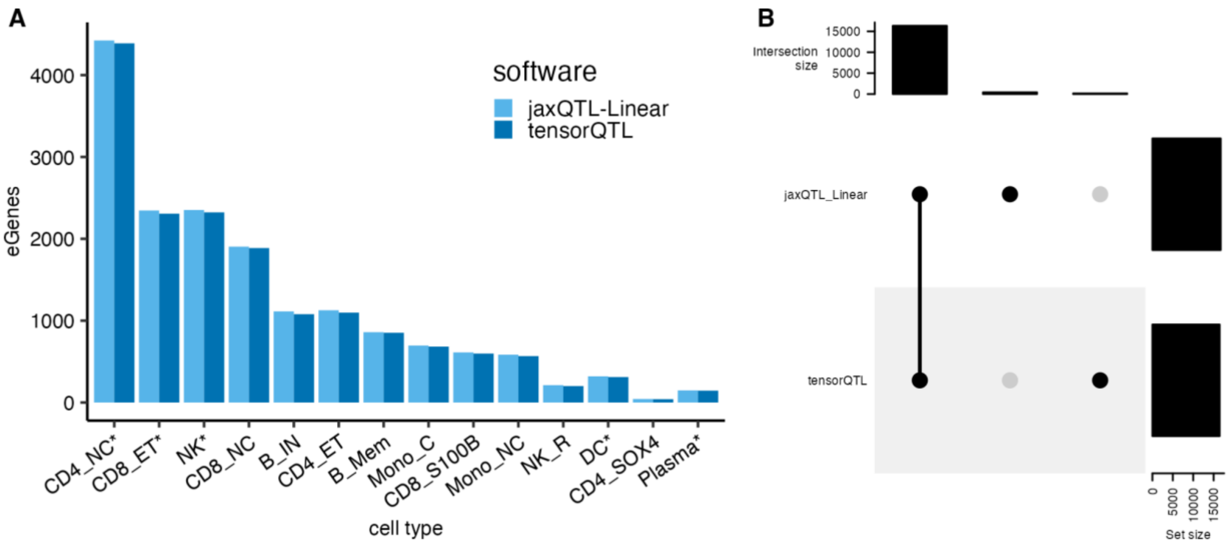


**Figure S14. eGene results are concordant between jaxQTL-Linear and tensorQTL.**
**(A)** We report the number of eGenes identified by jaxQTL-Linear and tensorQTL for genes expressed with sample-coverage > 10% of individuals. **(B)** We report eGenes overlap between these two software in the upset plot. The difference is up to the gene-level *P* value obtained by the permutation approach.

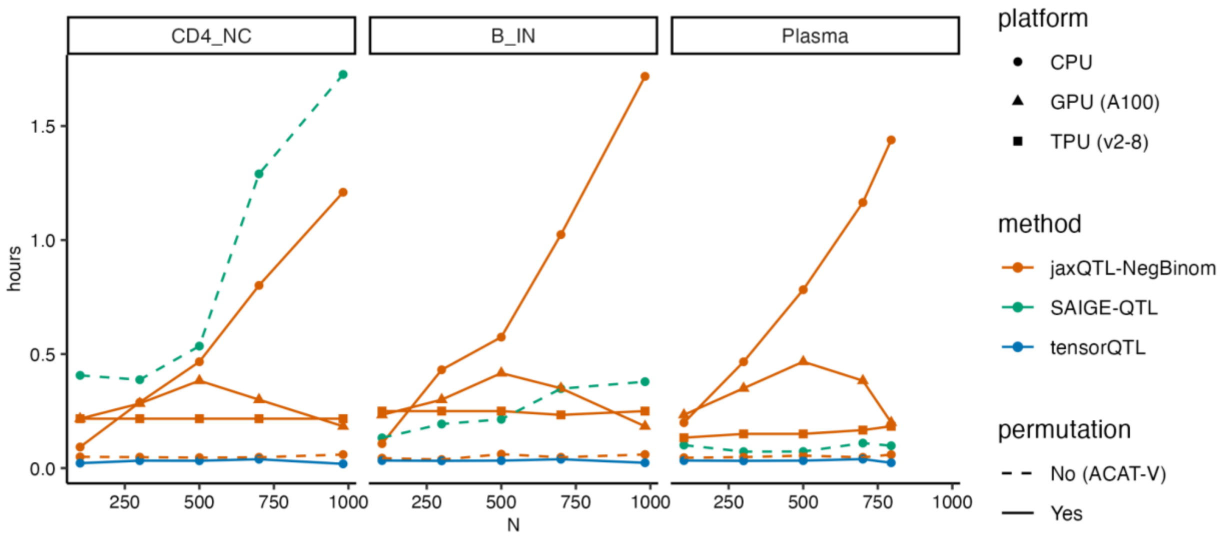


**Figure S15. Computational run time of sc-eQTL mapping on 50 randomly selected genes.** We report the runtime of cis-eQTL mapping across different software (jaxQTL-negbinom, SAIGE-QTL, or tensorQTL), platform (CPU, GPU, and TPU), and gene-level P value calculation approach (either permutation or ACAT-V method). We randomly selected 50 genes from chromosome 1 that are expressed in three representative cell types (CD4_NC, B_IN, Plasma) and created downsampling data for varying sample sizes (N = 100, 300, 500, 700).

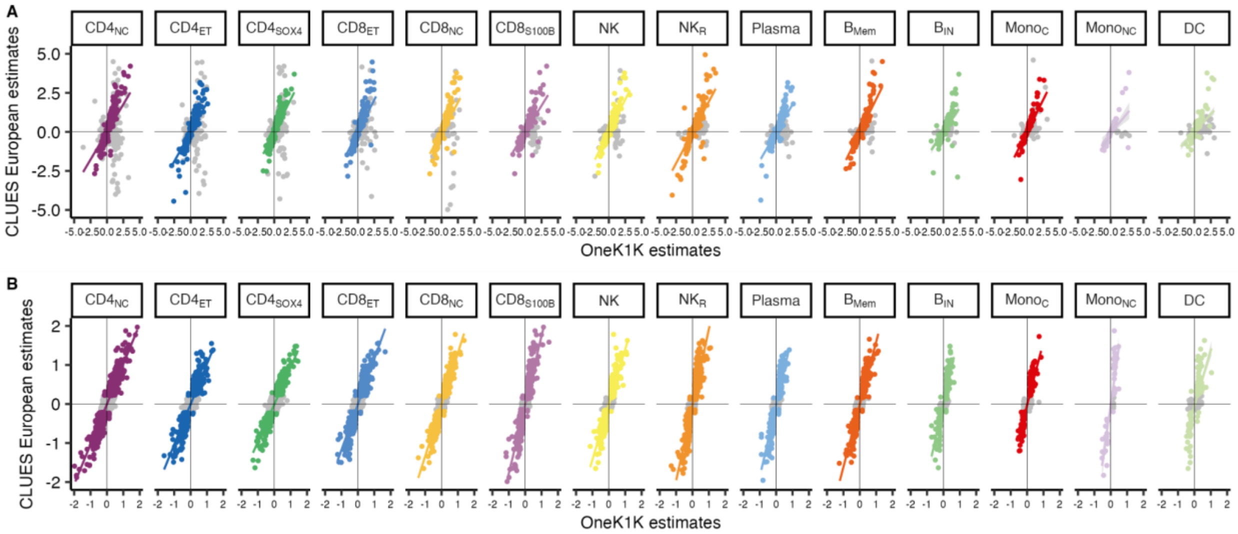


**Figure S16. Replication of sc-eQTLs effects in European samples from the CLUES study.** For 14,229 lead SNP-eGene pairs in their matched cell type from 88 European ancestry individuals in the CLUES study, we plot the raw slope estimate **(A)** and adjusted slope estimates **(B)** in CLUES versus OneK1K samples. Colored points are pairs replicated at FDR < 0.05 and grey points are null. 199 pairs with absolute raw slope > 5 in CLUES **(A)** and 35 pairs with absolute adjusted slope estimate > 2 **(B)** were truncated for visualization. The colored line is a fitted line with a 95% confidence band. (see full results in **Table S5**).

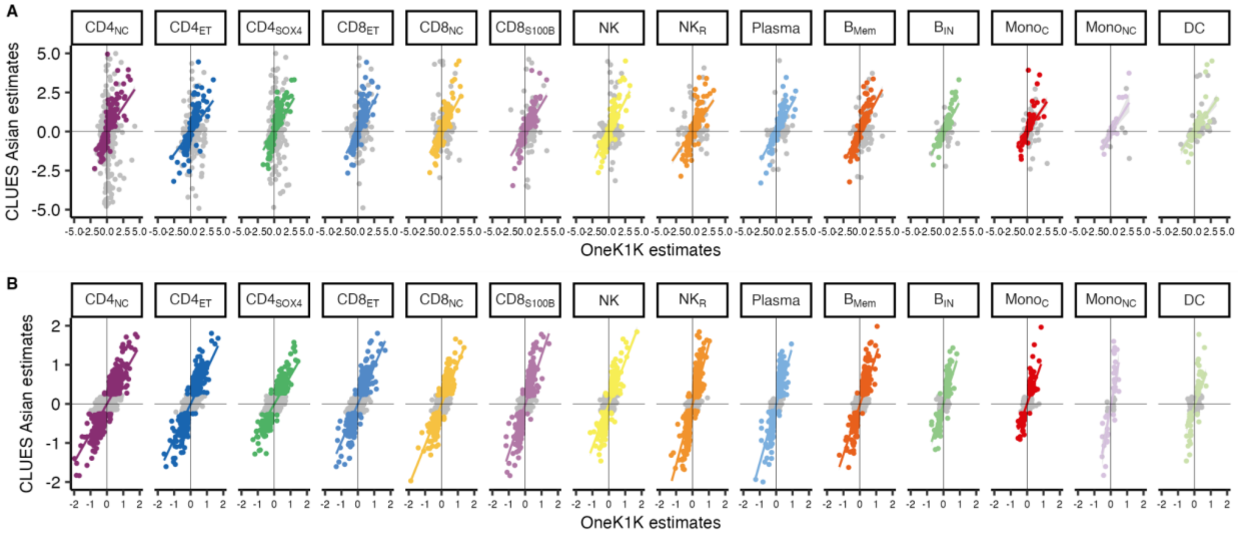


**Figure S17. Replication of sc-eQTLs effects in Asian samples from the CLUES study.** For 13,579 lead SNP-eGene pairs in their matched cell type from 88 Asian ancestry individuals in the CLUES study, we plot the raw slope estimate **(A)** and adjusted slope estimates **(B)** in CLUES versus OneK1K samples. Colored points are pairs replicated at FDR < 0.05 and grey points are null. 199 pairs with absolute raw slope > 5 in CLUES **(A)** and 35 pairs with absolute adjusted slope estimate > 2 **(B)** were truncated for visualization. The colored line is a fitted line with a 95% confidence band. (see full results in **Table S5**).

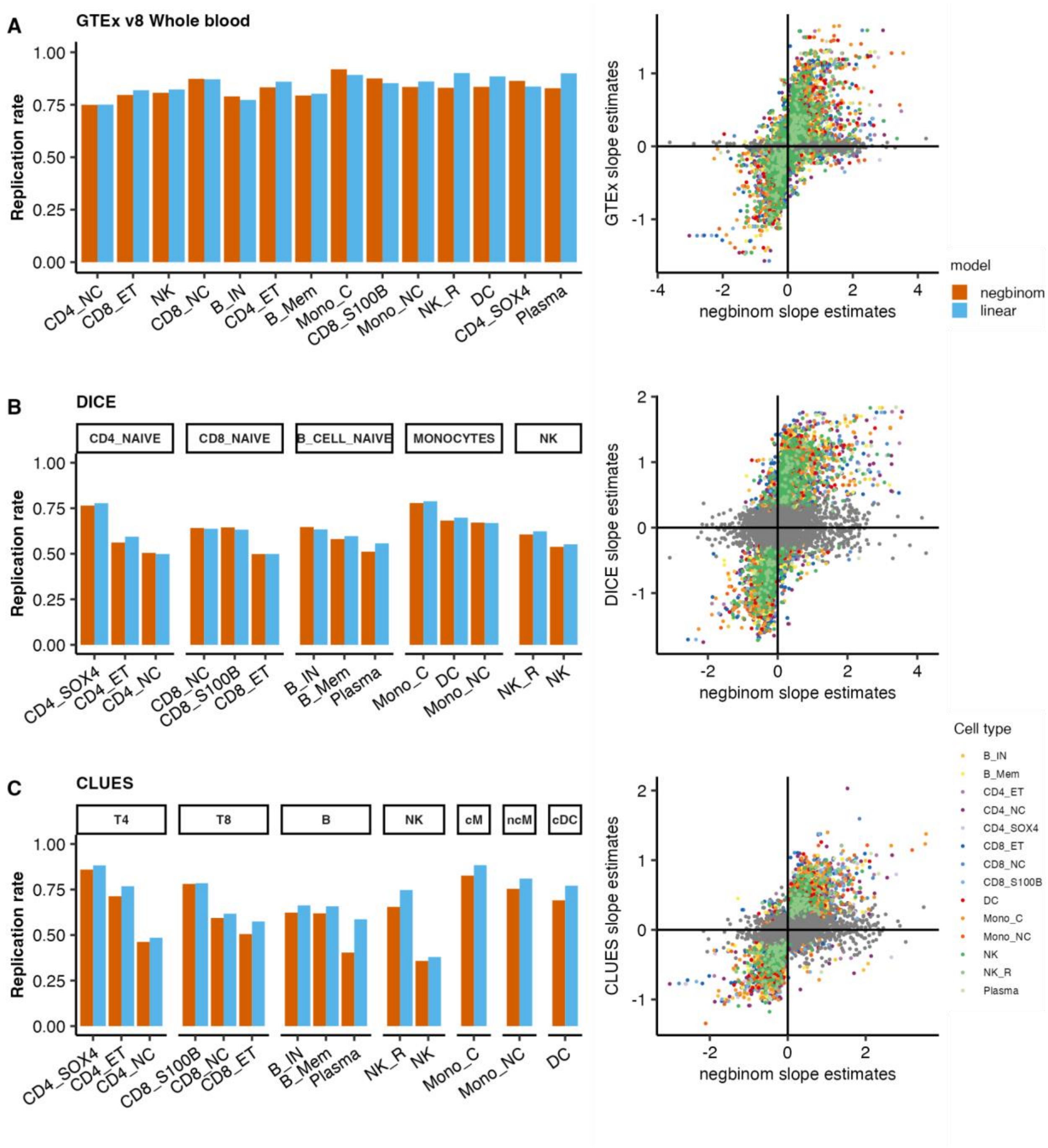


**Figure S18. Replication of lead SNP-eGene in tissue and cell-type level eQTLs.**
We report the replication rate at FDR < 0.05 of sc-eQTLs in matched cell types (tabs) identified by jaxQTL-negbinom and jaxQTL-linear in GTEx v8 EUR Whole blood **(A)**, DICE study **(B)**, CLUES cohort results calculated by eQTL catalogue **(C)**. For each replication cohort, we plot the raw slope estimates of these sc-eQTLs from jaxQTL-negbinom and the replication cohort. Colored points are pairs replicated at FDR < 0.05 and grey points are otherwise. T4: CD4+ T cells; T8: CD8+ T cells; NK: natural killer cells; cM: CD14+ conventional monocytes; ncM: CD16+ unconventional monocytes; cDC: conventional dendritic cells.

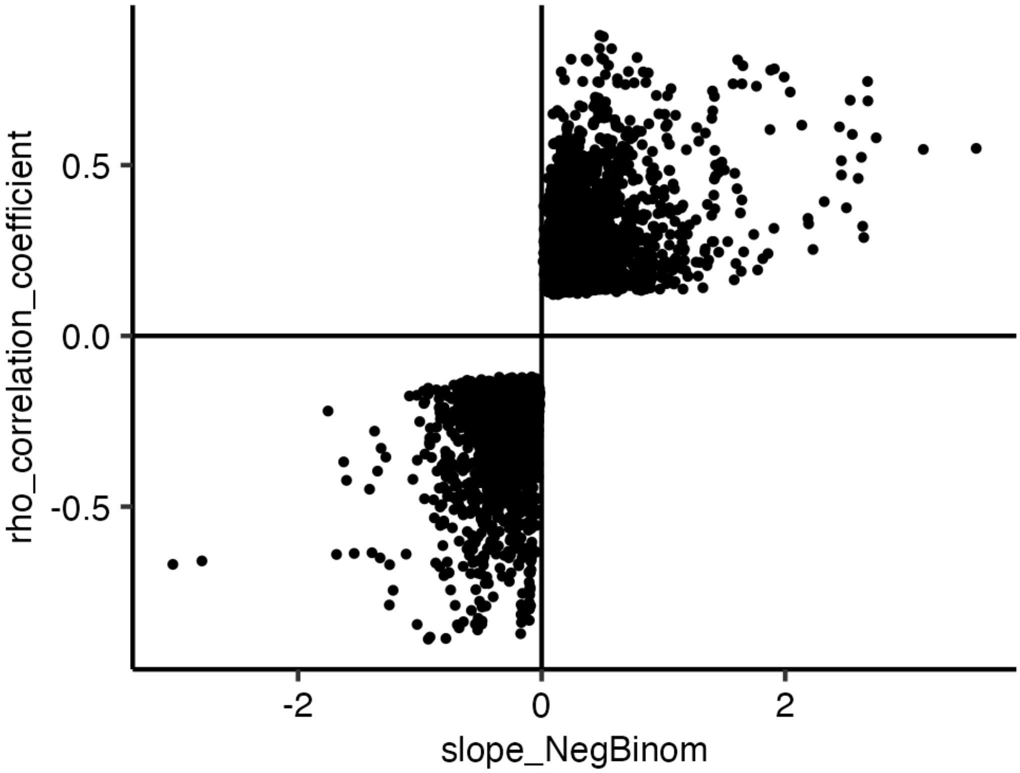


**Figure S19. Shared lead eSNP-eGene are consistent in direction between jaxQTL-negbinom and OneK1K linear model results.**

We report the effect size estimates obtained in jaxQTL-negbinom and rho correlation estimated in OneK1K results for 6,336 eQTLs identified in both studies across 14 cell types.

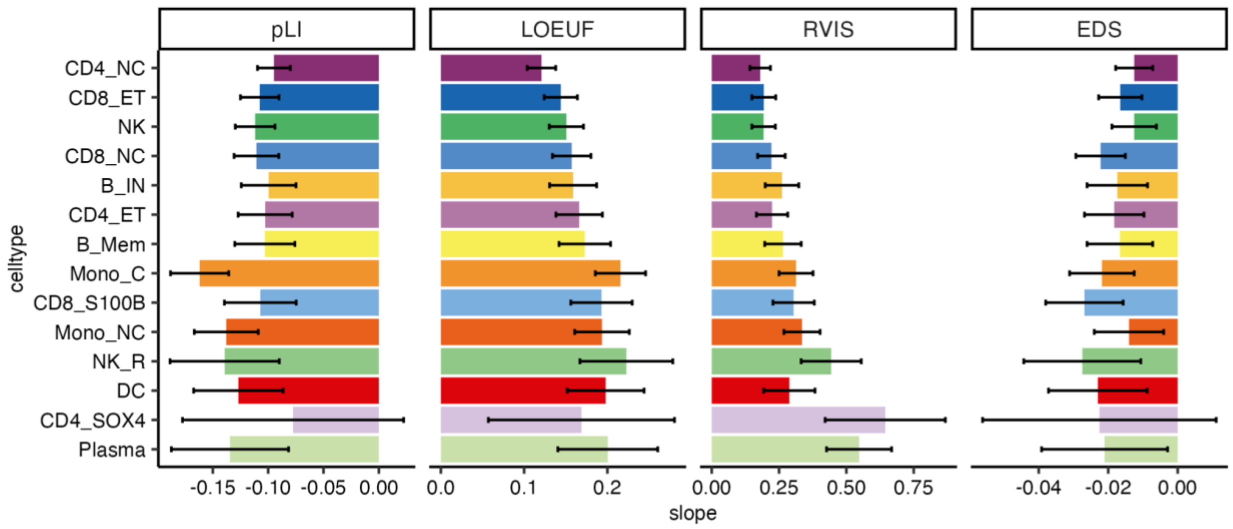


**Figure S20. eGenes are depleted of loss-of-function and enhancer domains.**
We report the slope of gene metrics on whether genes were identified as eGenes in each cell type after adjusting for gene expression. We report the fitted slope and error bars for 95% CI calculated by bootstrapping. Cell types were ordered by cell type proportion from high to low.

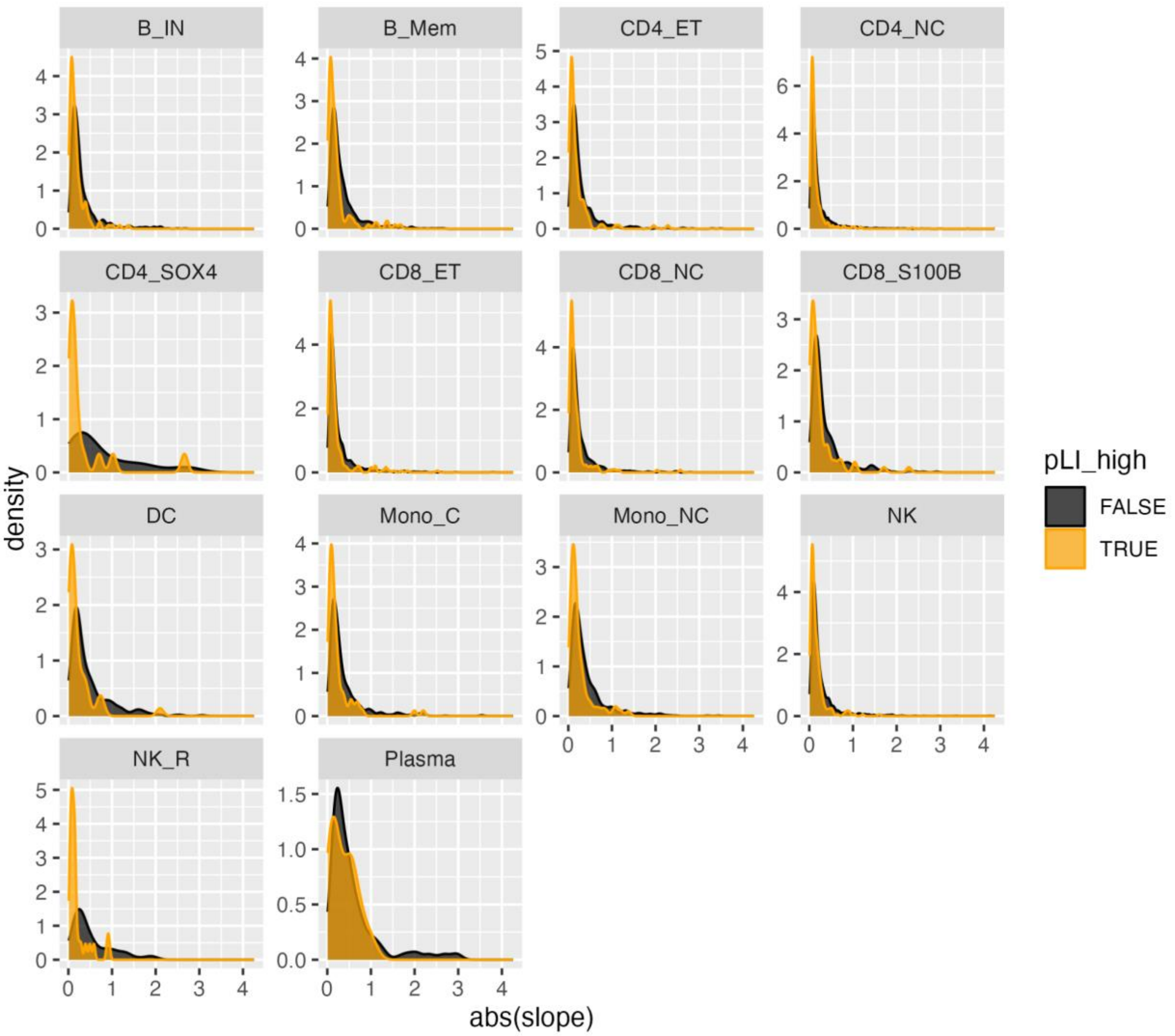


**Figure S21. eGenes depleted of loss-of-function have smaller sc-eQTL effect sizes.** We report the density of the absolute effect size of lead SNPs from eGenes, stratified by whether the eGene is depleted of loss-of-function (pLI > 0.9) or more tolerant (pLI < 0.9).

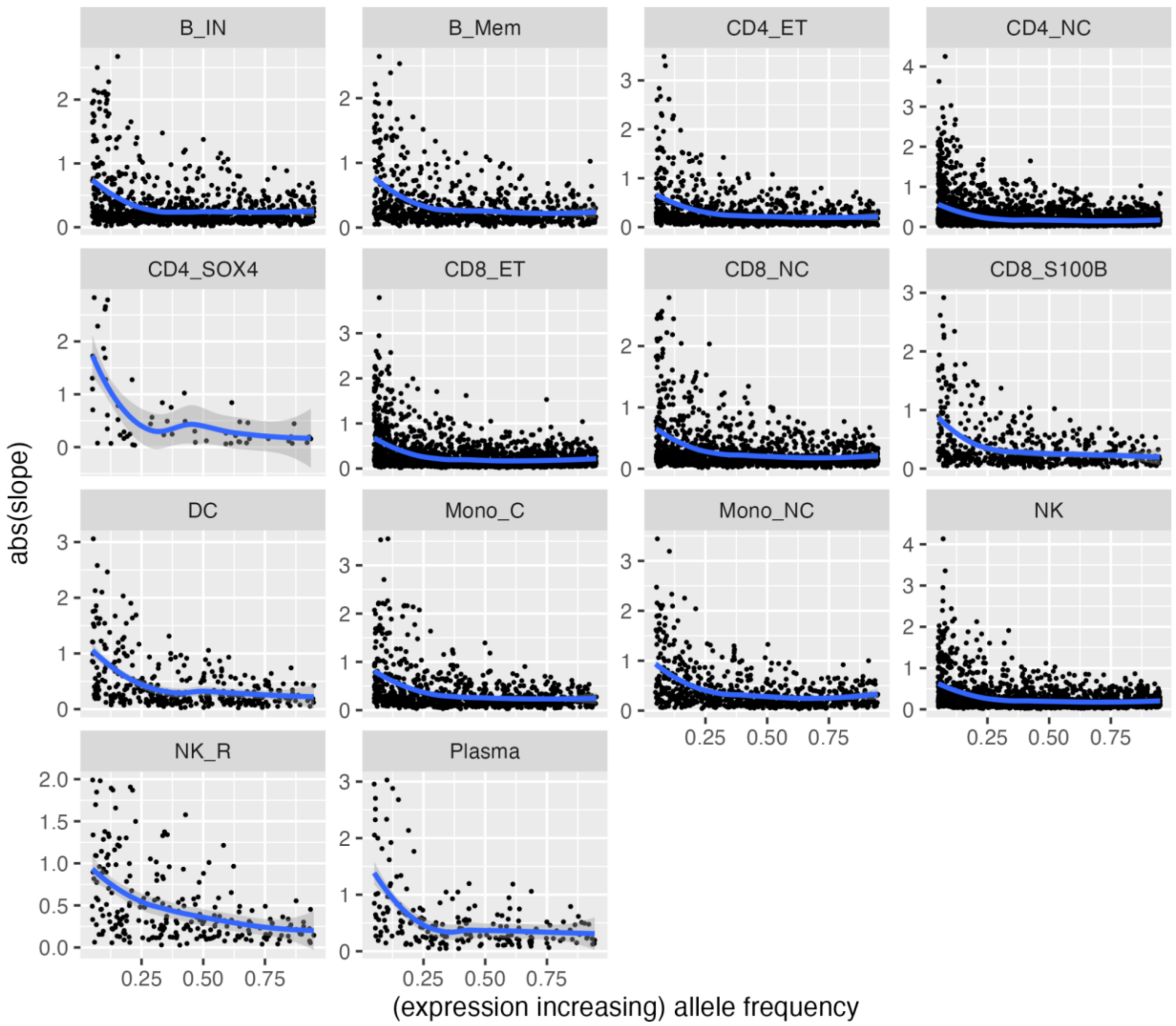


**Figure S22. Effect sizes of lead SNPs are greater at a lower allele frequency.**
We report the absolute slope as a function of allele frequency for alleles of the lead SNP eGenes that increase gene expression. The blue line and 95% CI confidence band (grey) were fitted by the Loess regression.

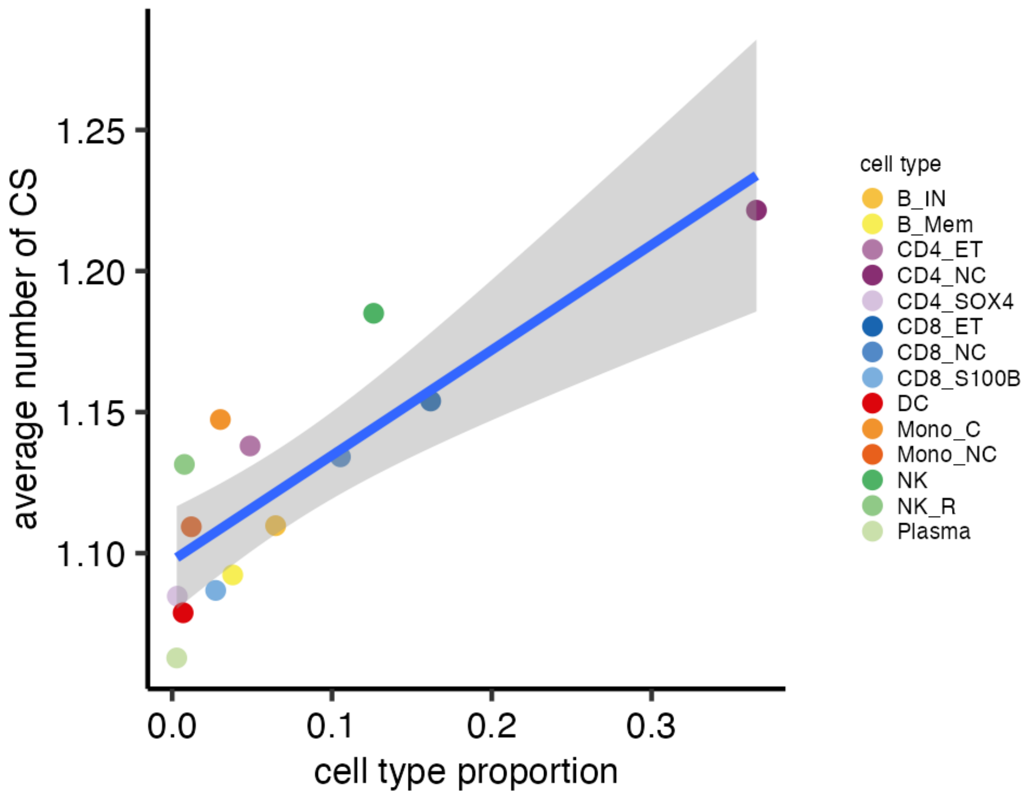


**Figure S23. The number of independent causal signals tracked with cell type proportions.** We report the average number of credible sets (CS) for each cell type as a function of cell type proportion. The blue line is fitted with linear regression and 95% CI in grey.

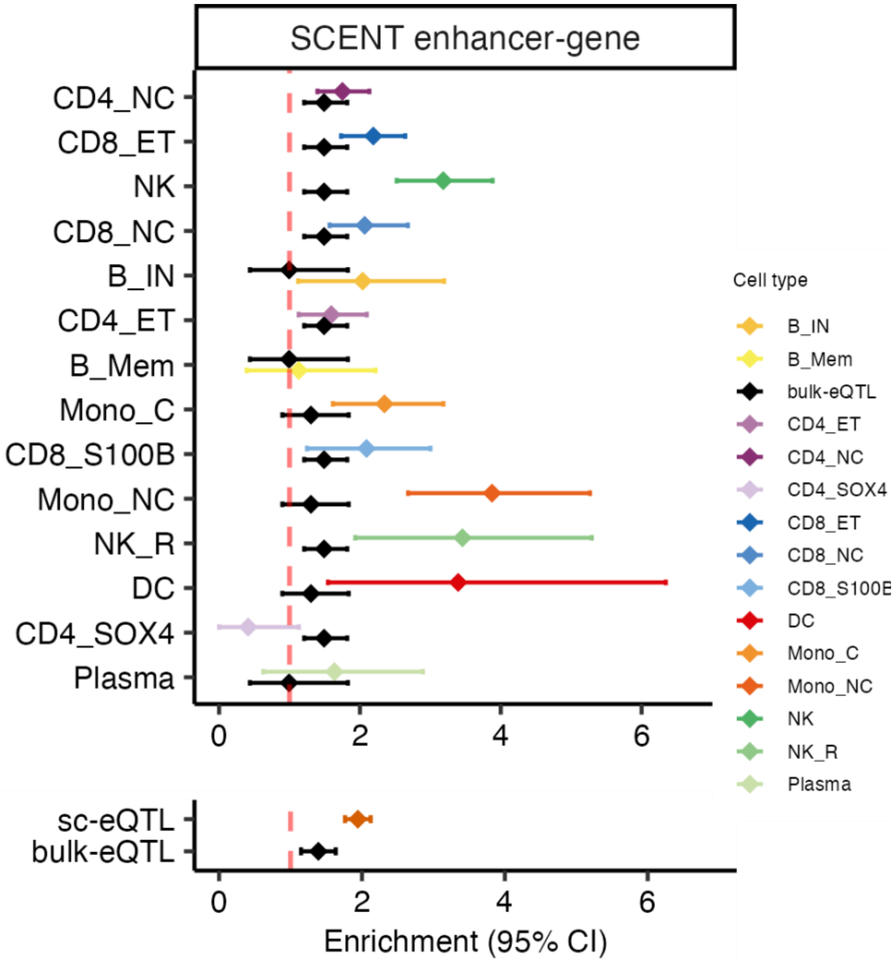


**Figure S24. sc-eQTLs are more enriched in enhancer-gene links identified by SCENT.**
We report the enrichment in sc-eQTLs (matched by SCENT cell type) and bulk-eQTL for enhancer-gene link identified by the SCENT approach respectively. We report a meta-analysis across 14 cell types at the bottom.

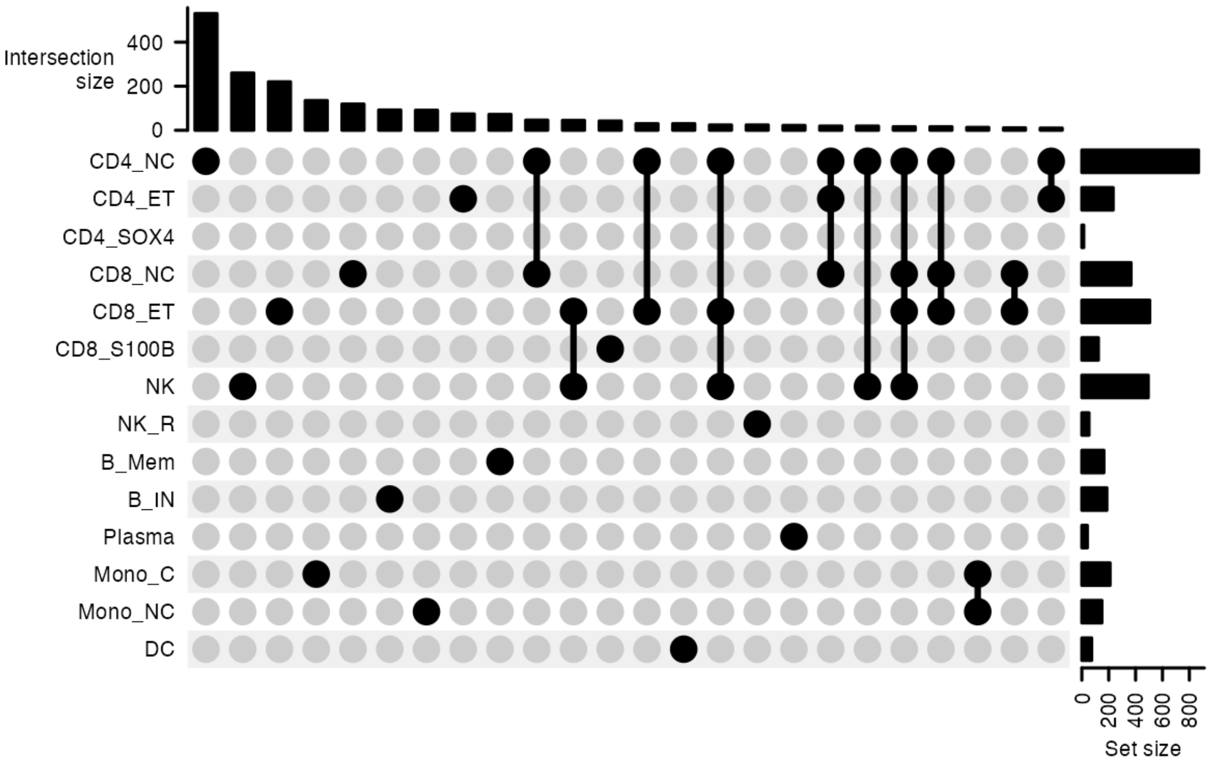


**Figure S25. sc-eQTL sharing by simple counting approach.**

For 2,256 sc-eQTL with PIP > 0.5 in at least one cell type, we report sc-eQTL sharing by magnitude in an UpSet plot, where bars in upper panels denote the number of sc-eQTLs found exclusively in a given intersection set for connected dots. Bars on the right side are the number of sc-eQTLs shared or specific in each cell type.

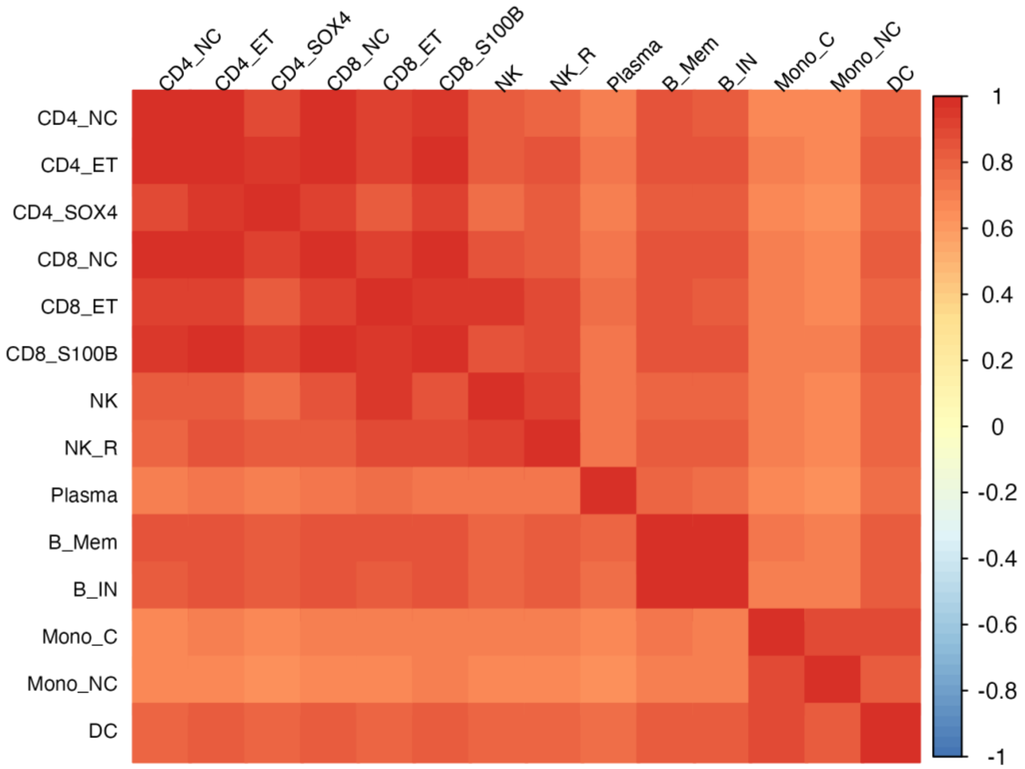


**Figure S26. Correlation matrix for the weighted sum of data-driven covariance matrices.** After fitting *mashr* on a random set of 21,542 eQTLs using 27 covariance matrices learned from 2,256 fine-mapped sc-eQTLs, we took the weighted sum of these covariance matrices and plotted a heatmap of its correlation matrix.

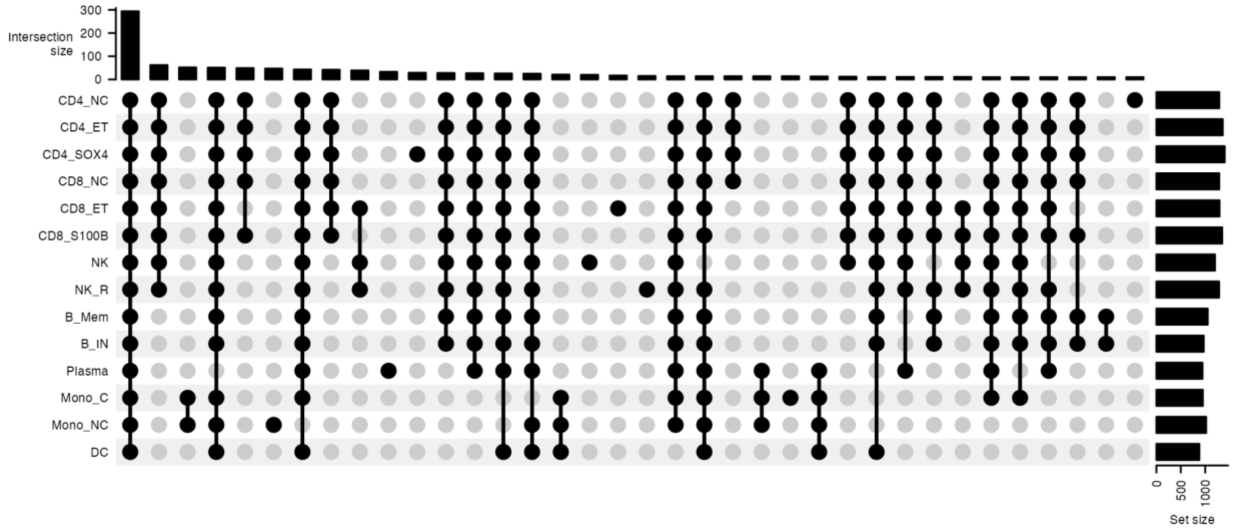


**Figure S27. sc-eQTL sharing by magnitude.**

For 2,012 sc-eQTL with LFSR < 0.05 in at least one cell type, we report sc-eQTL sharing by magnitude in an UpSet plot, where bars in upper panels denote the number of sc-eQTLs found exclusively in a given intersection set for connected dots. Bars on the right side are the number of sc-eQTLs shared or specific in each cell type.

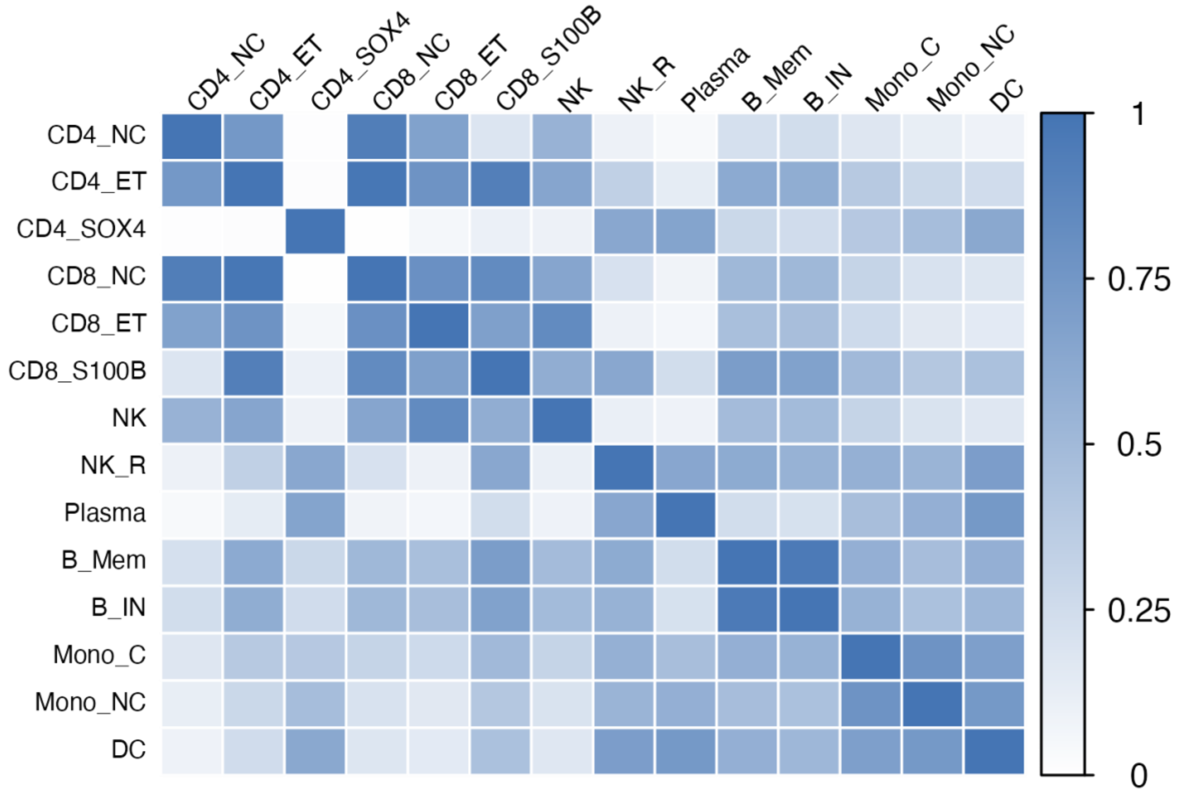


**Figure S28. Similar cell types share eQTL by magnitude.**
We report *mashr* pairwise sharing by magnitude between cell types, which is restricted to significant eQTL at LFSR < 0.05 in at least one of the cell types.

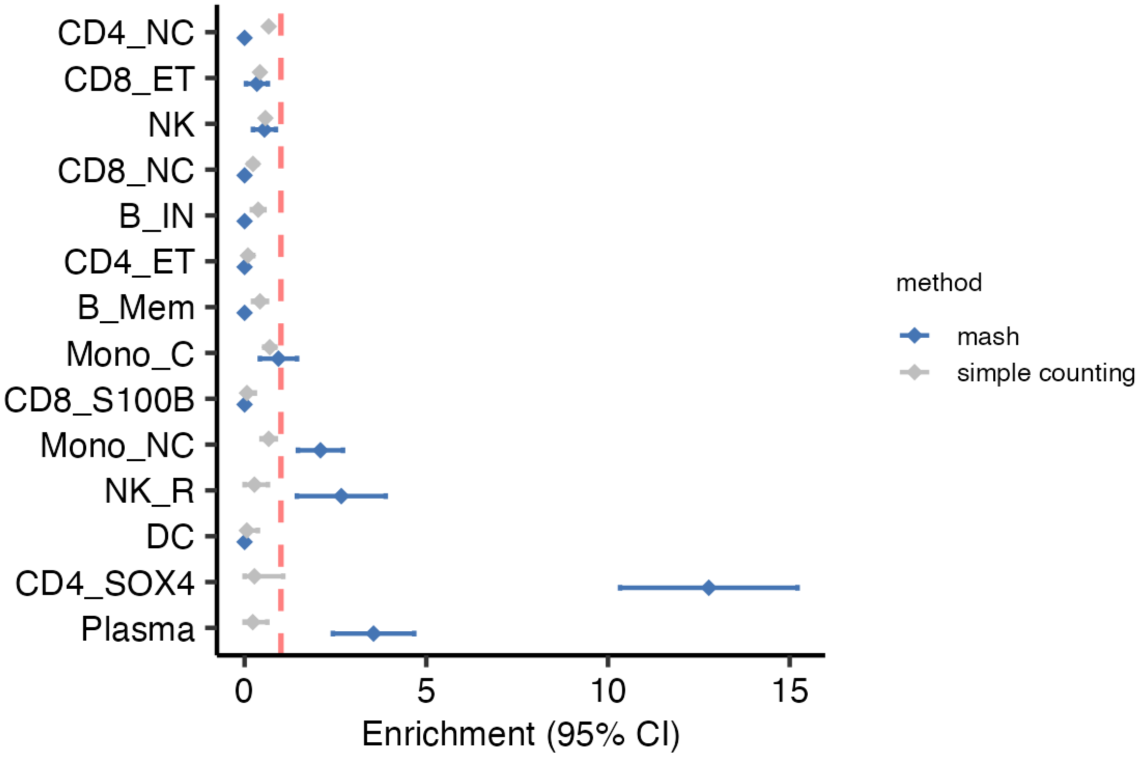


**Figure S29. Enrichment of cell-type-specific open chromatins in cell-type-specific sc-eQTLs.**
We report the enrichment of ATAC-seq peaks exclusive to each cell type in cell-type-specific sc-eQTLs identified by *mashr* or simple counting approach (see **Methods**). The standard errors were obtained by permutations on cell type labels.

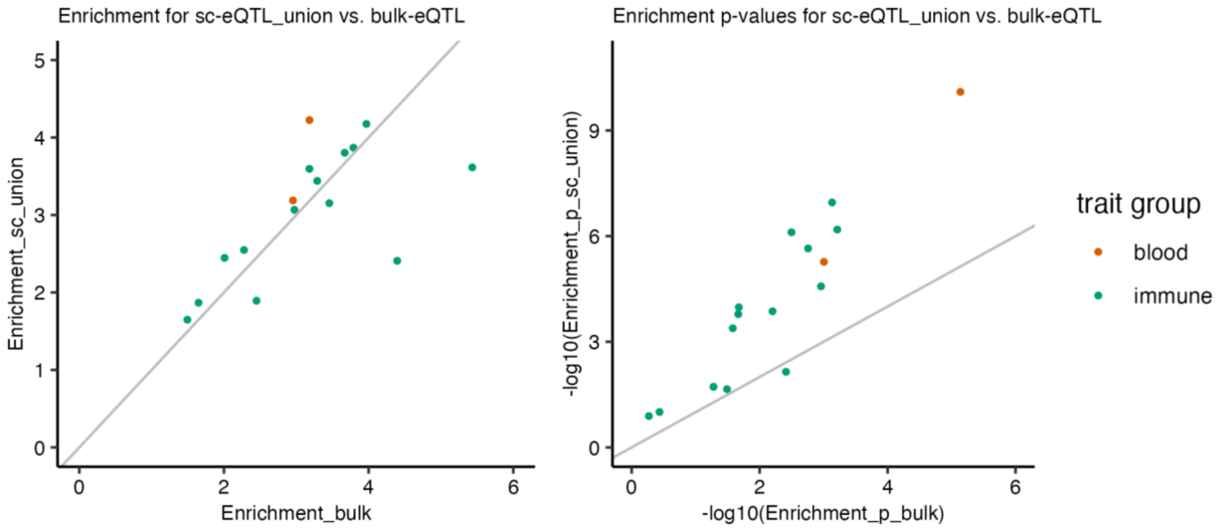


**Figure S30. Enrichment of heritability in GWAS traits for sc-eQTL_union and bulk-eQTLs obtained by S-LDSC is comparable.**

We report the heritability enrichment **(A)** and corresponding enrichment p-values **(B)** of sc-eQTL (union) and bulk-eQTL annotations for 16 GWAS immune traits. Traits were categorized and colored by blood and immune disease groups.

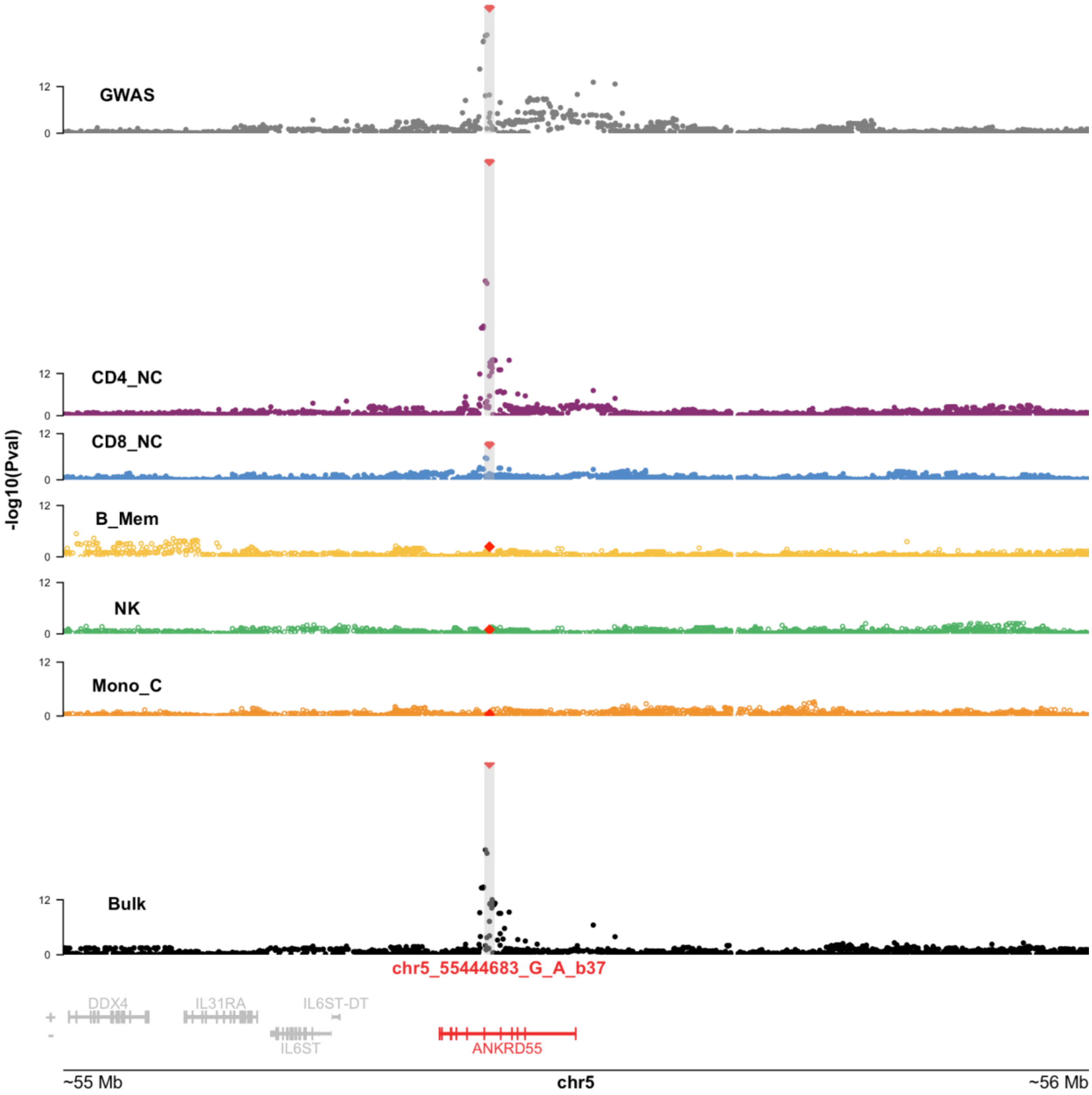


**Figure S31. Colocalization of *ANKRD55* eQTLs with RA GWAS results.**
Figure similar to Figure 6 but reporting *ANKRD55* rather than *IL6ST* eQTLs.

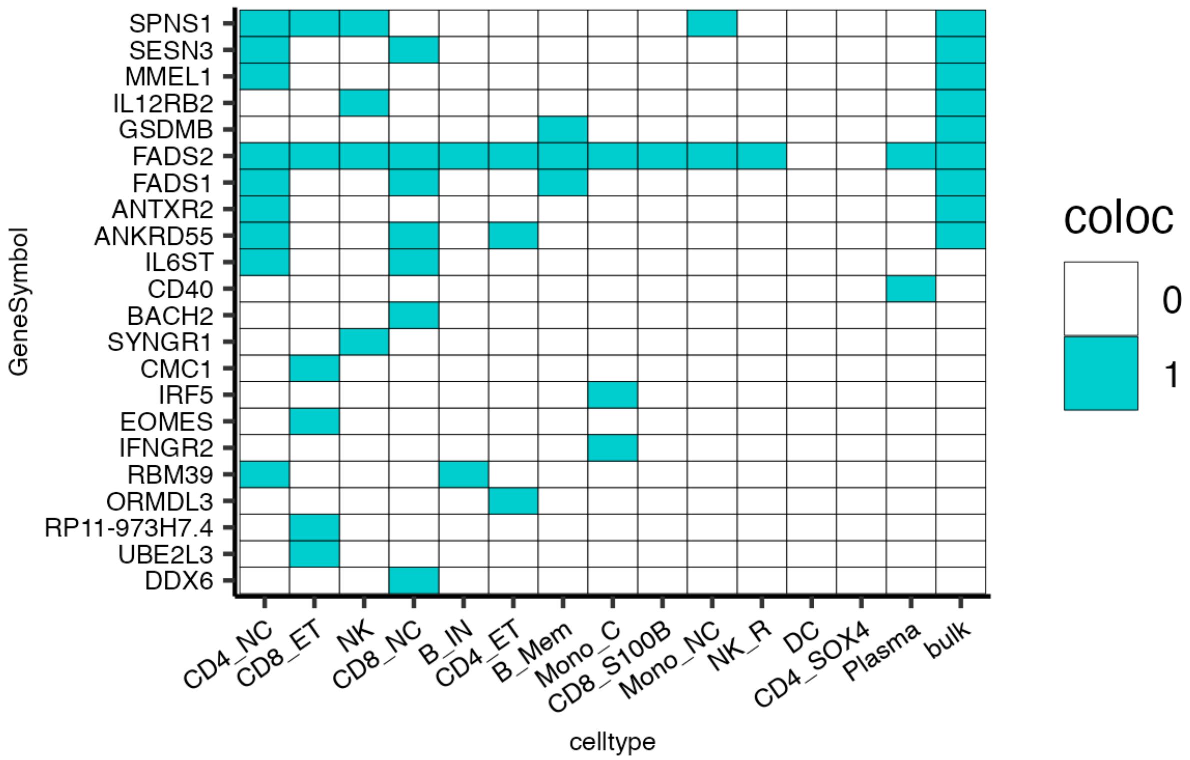


**Figure S32. Colocalization of sc-eQTL and GWAS results for RA.**
We report colocalization results between RA GWAS and OneK1K sc-eQTLs. Each blue-colored square represents a colocalization signal (PP.H4 > 0.9) for a gene (row) in a given cell type (column).
