## supplemental note for "Efficient count-based models improve power and robustness for large-scale single-cell eQTL mapping"

### Supplemental note for jaxQTL

This math note aims to show structural similarity between the Poisson mixed effect (PME) model and the negative binomial (negbinom) model under specific assumptions.

#### 1 Count-based models for read counts

##### 1.1 Poisson Mixed Effect model

For a focal gene, let $y_{ij}$ be the count of aligned reads from the $j$th cell for the $i$th individual observed in scRNA-seq data and let $\mathbf{x}_i^\intercal$ be their vector of $k$ covariates. To incorporate library size per cell, we add the offset term $l_{ij}$ in the mean component, where $l_{ij} := \sum_g l_{ijg}$ across genes. Using a log link function, the Poisson mixed effect (PME) model for single-cell count can be written as:

$$\begin{aligned} y_{ij} \mid u_i &\sim \text{Pois}(\lambda_{ij}) \\ \lambda_{ij} &:= \exp(\mathbf{x}_i^\intercal \boldsymbol{\beta} + u_i + \log l_{ij}) \\ u_i &\sim \mathcal{N}(0, \sigma_u^2), \end{aligned}$$

where $u_i$ is the random intercept for the $i$th individual which accounts for intra-individual correlation between cells. Here we assume individuals are unrelated.

Next, we define pseudobulk-level count for the same focal gene in the $i$th individual by $y_i = \sum_{j=1}^{m_i} y_{ij}$, where $m_i$ denote the number of cells for the $i$th individual. Similary, the pseudobulk-level library size becomes $l_i := \sum_j l_{ij}$.

##### 1.2 Negative binomial model

Using the definitions above, we can model the psuedobulk counts directly. Namely, using a log link function, the negative binomial model of pseudobulk $y_i$ can be represented by,

$$\begin{aligned} y_i &\sim \text{NB}(\lambda_i, \alpha) \\ \lambda_i &:= \exp(\mathbf{x}_i^\intercal \boldsymbol{\beta} + \log l_i), \end{aligned}$$

where $\mathbb{E}[y_i] = \lambda_i$ and $\alpha$ models overdispersion for the variance given by $\mathbb{V}[y_i] = \lambda_i + \alpha\lambda_i^2$. When $\alpha = 0$, the negbinom model reduces to the Poisson model.

#### 2 Structural Similarity of Variance

Here we demonstrate the structural similarity of the two models up to the second central-moment when considering psuedobulk count data. We will make clear the base distribution when deriving expectations and variances, unless unambiguous by the context.

#### 2.1 PME Marginal Mean

We start with establishing the mean relationship between pseudobulk count and single-cell count based on PME model. By linearity, the marginal expectation of pseudobulk count $\mathbb{E}[y_i]$ can be calculated as a sum of expected single-cell count in PME model as,

$$\mathbb{E}[y_i] = \mathbb{E}\left[\sum_j^{m_i} y_{ij}\right] = \sum_j^{m_i} \mathbb{E}[y_{ij}].$$

The marginal expectation of single-cell count $\mathbb{E}[y_{ij}]$ in PME model is calculated by,

$$\begin{aligned}
\mathbb{E}[y_{ij}] &= \mathbb{E}[\mathbb{E}[y_{ij} \mid u_i]] \\
&= \mathbb{E}[\exp(\mathbf{x}_i^\intercal \boldsymbol{\beta} + u_i + \log l_{ij})] \\
&= \exp(\mathbf{x}_i^\intercal \boldsymbol{\beta} + \log l_{ij}) \mathbb{E}[\exp(u_i)] \\
&= \exp(\mathbf{x}_i^\intercal \boldsymbol{\beta} + \log l_{ij}) \exp(\sigma_u^2/2),
\end{aligned}$$

where $\mathbb{E}[\exp(u_i)] = \exp(\sigma_u^2/2)$ is the moment generating function of $u_i$ evaluated at $t = 1$. It follows that the pseudobulk mean becomes,

$$\begin{aligned}
\mathbb{E}[y_i] = \mathbb{E}[\sum_j^{m_i} y_{ij}] &= \sum_j^{m_i} \mathbb{E}[y_{ij}] \\
&= \sum_j^{m_i} \exp(\mathbf{x}_i^\intercal \boldsymbol{\beta} + \log l_{ij}) \exp(\sigma_u^2/2) \\
&= \exp(\mathbf{x}_i^\intercal \boldsymbol{\beta} + \sigma_u^2/2) \sum_j^{m_i} \exp(\log l_{ij}) \\
&= \exp(\mathbf{x}_i^\intercal \boldsymbol{\beta} + \sigma_u^2/2) \sum_j^{m_i} l_{ij} \\
&= \exp(\mathbf{x}_i^\intercal \boldsymbol{\beta} + \sigma_u^2/2) l_i \\
&= \exp(\mathbf{x}_i^\intercal \boldsymbol{\beta} + \log l_i) \exp(\sigma_u^2/2).
\end{aligned}$$

We observe that the per-cell library size naturally collapses to individual library size in $l_i = \sum_j^{m_i} l_{ij}$.

When $\sigma_u^2 \to 0$, the mean component of PME model approaches the standard Poisson model.

#### 2.2 PME Marginal Variance

The marginal variance of pseudobulk count $\mathbb{V}[y_i]$ under the PME model can be derived in a similar fashion by using the law of total variance as,

$$\mathbb{V}[y_i] = \mathbb{V}[\mathbb{E}[y_i \mid u_i]] + \mathbb{E}[\mathbb{V}[y_i \mid u_i]].$$

The first term is expanded as,

$$\begin{aligned}
\mathbb{V}[\mathbb{E}[y_i \mid u_i]] = \mathbb{V}[\mathbb{E}[\sum_j^{m_i} y_{ij} \mid u_i]] &= \mathbb{V}[\exp(u_i)] \left[\sum_j^{m_i} \exp(\mathbf{x}_i^\intercal \boldsymbol{\beta} + \log l_{ij})\right]^2 \\
&= (\exp(2\sigma_u^2) - \exp(\sigma_u^2)) \left[\sum_j^{m_i} \exp(\mathbf{x}_i^\intercal \boldsymbol{\beta} + \log l_{ij})\right]^2 \\
&= (\exp(2\sigma_u^2) - \exp(\sigma_u^2)) \exp(\mathbf{x}_i^\intercal \boldsymbol{\beta} + \log l_i)^2.
\end{aligned}$$

Recall the per-cell counts $y_{ij}$ are conditionally independent given $u_i$ and that the conditional variance $\mathbb{V}[y_{ij} \mid u_i] = \mathbb{E}[y_{ij} \mid u_i]$ under the PME model. Noting this we can expand the second term as,

$$\begin{aligned}
\mathbb{E}[\mathbb{V}[y_i \mid u_i]] &= \mathbb{E}[\mathbb{V}[\sum_j^{m_i} y_{ij} \mid u_i]] = \mathbb{E}[\sum_j^{m_i} \mathbb{V}[y_{ij} \mid u_i]] \\
&= \mathbb{E}[\sum_j^{m_i} \mathbb{E}[y_{ij} \mid u_i]] = \sum_j^{m_i} \mathbb{E}[\mathbb{E}[y_{ij} \mid u_i]] \\
&= \sum_j^{m_j} \mathbb{E}[y_{ij}] = \exp(\mathbf{x}_i^\intercal \boldsymbol{\beta} + \log l_i) \exp(\sigma_u^2/2).
\end{aligned}$$

Finally, the marginal variance becomes:

$$\begin{aligned}
\mathbb{V}[y_i] &= \underbrace{(\exp(2\sigma_u^2) - \exp(\sigma_u^2)) \exp(\mathbf{x}_i^\intercal \boldsymbol{\beta} + \log l_i)^2}_{\mathbb{V}[\mathbb{E}[y_i \mid u_i]]} + \underbrace{\exp(\mathbf{x}_i^\intercal \boldsymbol{\beta} + \log l_i) \exp(\sigma_u^2/2)}_{\mathbb{E}[\mathbb{V}[y_i \mid u_i]]} \\
&= (\exp(\sigma_u^2) - 1) \exp(\mathbf{x}_i^\intercal \boldsymbol{\beta} + \log l_i)^2 \exp(\sigma_u^2) + \exp(\mathbf{x}_i^\intercal \boldsymbol{\beta} + \log l_i) \exp(\sigma_u^2/2) \\
&= (\exp(\sigma_u^2) - 1) \left[\exp(\mathbf{x}_i^\intercal \boldsymbol{\beta} + \log l_i) \exp(\sigma_u^2/2)\right]^2 + \exp(\mathbf{x}_i^\intercal \boldsymbol{\beta} + \log l_i) \exp(\sigma_u^2/2) \\
&= (\exp(\sigma_u^2) - 1)\mathbb{E}[y_i]^2 + \mathbb{E}[y_i] \\
&= \mathbb{E}[y_i] + (\exp(\sigma_u^2) - 1)\mathbb{E}[y_i]^2.
\end{aligned}$$

Again, when $\sigma_u^2 \to 0$, the marginal variance of the PME model approaches the standard Poisson model.

#### 2.3 Structural Equivalence of Moments

Recall the (marginal) means for $y_i$ under each distribution given by,

$$\begin{aligned}
\mathbb{E}_{\text{PME}}[y_i] &= \exp(\sigma_u^2/2) \exp(\mathbf{x}_i^\intercal \boldsymbol{\beta} + \log l_i) \\
\mathbb{E}_{\text{NB}}[y_i] &= \exp(\mathbf{x}_i^\intercal \boldsymbol{\beta} + \log l_i).
\end{aligned}$$

When $\sigma_u^2$ is small $\exp(\sigma_u^2/2) \approx 1$, thus $\mathbb{E}_{\text{PME}}[y_i] \approx \mathbb{E}_{\text{NB}}[y_i]$, which becomes exact when $\sigma_u^2 \to 0$.

Similarly for the (marginal) variance of $y_i$ we have,

$$\begin{aligned}
\mathbb{V}_{\text{PME}}[y_i] &= \mathbb{E}_{\text{PME}}[y_i] + (\exp(\sigma_u^2) - 1)\mathbb{E}_{\text{PME}}[y_i]^2 \\
\mathbb{V}_{\text{NB}}[y_i] &= \mathbb{E}_{\text{NB}}[y_i] + \alpha \mathbb{E}_{\text{NB}}[y_i]^2,
\end{aligned}$$

which demonstrates a structural similarity between models where the variance is given by $\mu + c\mu^2$ with respective means $\mu$ and adjustment term $c$. As $c \to 0$ both models reduce to the standard Poisson model.
